# The contribution of the out-of-home food (OOHF) sector to the national diet: a cross-sectional survey with repeated 24-hour recalls of adults in England (2023-2024)

**DOI:** 10.1101/2025.06.30.25330369

**Authors:** James Garbutt, Nick Townsend, Laura Johnson, Andrew Jones, Martin O’Flaherty, Zoé Colombet, Amy Finlay, Eric Robinson, Zoi Toumpakari

## Abstract

**Background:** Quantifying the contribution of the out-of-home food and drink (OOHF) sector on the English national diet could inform effective nutrition policy development. However, rigorous data and analysis for estimating the relative contribution of the OOHF sector to total diet is lacking.

**Objectives:** To quantify the nutritional contribution per day and per eating occasion of the OOHF sector on adults’ diets in England in terms of energy, total fat, saturated fat, sugar and salt, overall and across key demographics.

**Design:** Data collected from n=1232 participants (51% female) in a multi-stage online survey including demographics and mean 4.5 (SE 0.03) 24-hr dietary recalls over a 2-3 week period. Four waves of data collection (September 2023 -May 2024) captured seasonal differences. Hierarchical multilevel models explored differences in energy between 1) eating occasions or 2) days, containing OOHF versus not. Total nutritional differences by OOHF across demographics were described.

**Results:** Median out-of-home (OOH) eating occasion energy intakes were 440kcal (vs 294kcal for non-OOH occasions). After adjustment, energy intakes during OOH eating occasions were 196kcal [95%CI: 171, 221] (p<0.001) larger than non-OOH occasions. OOH days contained 103kcal [29, 177] (p=0.006) more than non-OOH days, but evidence of association was not robust to multiple testing. OOHF contributed a median of 11.0% (IQR: 0, 23.4) energy to total weekly energy intake. Participants exceeded recommended daily nutritional intakes more often on OOH days versus non-OOH days (>70g fat (female): 43% vs 29%, >90g fat (male): 29% vs 16%; >20g saturated fat (female): 62% vs 53%, >30g saturated fat (male): 36% vs 28%; >90g total sugars: 33% vs 26%; >6g salt: 32% vs 18%).

**Conclusions:** The OOHF sector makes a substantial contribution to the national English diet, with food prepared OOH linked to higher energy and poorer nutritional intakes. Population-wide nutrition policies targeting the sector are needed.

## Introduction

In the UK in 2021, 26% of adults were living with obesity [1] with the food environment identified as an important contributor to the obesity epidemic. A key aspect of this obesogenic food environment is the out-of-home food and drink (OOHF) sector, as frequent use is linked to poorer diet and weight gain [2–4]. The definition of the OOHF sector varies substantially in the literature, with some studies defining it as the source and place of food preparation [2–6] and others as the place of food consumption [2, 3, 7]. The latter, however, is likely to underestimate the true contribution of OOHF in people’s diets as it can exclude ready-to-eat and take-away meals eaten at home but prepared elsewhere. Despite the variability in OOHF definition, over the last 20 years, frequency of OOHF consumption has generally increased [8, 9], except for a temporary drop during COVID-19 [10]. Given its substantial and growing role on diet [2, 3], policies that target the OOHF sector have been identified as being key to reducing diet-related disease and obesity [11, 12].

To identify effective and equitable OOHF sector policies, it must first be understood what the exact contribution that OOHF sources have on a population’s diet. So far, the contribution of the OOHF sector on diet has been assessed at an individual or household level. At the individual level, out-of-home (OOH) eating is typically assessed via measuring frequency of eating out, without examining foods’ nutritional quality [8, 9, 13, 14], or via assessing daily OOH energy intake based on a single day’s food intake [4]. Given that greater variation in food intake exists within rather than between people (90% vs 10% respectively [15]), such methods cannot accurately capture the contribution of the OOHF sector. Additionally, while person-level methods provide information on daily food and nutrient intakes and associations with diet or health, confounding by person-level characteristics can’t be ruled out, which may overestimate of the contribution of OOHF specifically to total diet. We therefore do not know if food eaten in the OOHF sector is what impacts on poor diet/health or whether individuals who eat more OOHF are simply more likely to eat food of poor nutritional quality at other times too [16, 17]. For example, cross-sectional evidence from NHANES in the US, suggests that adults who are frequent fast-food consumers (n=4,012) have similar energy and nutrient intakes on days visiting a fast-food outlet compared to days when they do not [17]. In the UK, the National Diet and Nutrition Survey (NDNS), which, until 2018 only collected information on location of food consumption, has recently collected information on where food was bought or obtained. An unpublished government report reported that from 2019 to 2023 the OOHF sector contributed a mean of 12% to total energy intake [6]. However, NDNS nutritional data are matched to a limited database that does not have OOH-outlet-specific and up-to-date nutritional information on products sold in the OOHF sector. This makes NDNS unsuitable, especially as recent research reveals that the nutritional profile of similar product types (e.g. ‘a burger and fries meal’) varies dramatically (e.g. energy content range of ∼700kcals to ∼2000kcals) dependent on which outlet it is purchased from [18, 19].

At the household level, food purchasing data in the UK has also been used to estimate the contribution of OOHF [5, 20]. For example, evidence up to 2021 using Kantar Worldpanel Take Home and Out of Home purchasing data for 5,800 individuals estimated that the OOHF sector contributes an average of 300 kcal per person per day [5]. However, as individual OOHF purchases were accompanied by household, but not individual, at-home food data, it’s unclear to what extent OOHF added to or replaced non-OOHF intakes. It is possible that on days when individuals eat OOH, they eat less food at-home, in preparation or response to a large meal eaten out. However, it is currently unclear how OOHF purchases contribute to individual total dietary intakes. To estimate potential compensatory changes to food intake at-home in response to food eaten OOH, multiple days of individual food intakes are necessary. We also know that greater variation in food intake exists within individuals (between eating occasions) rather than between individuals (90% vs 10% respectively [15]), thus relying on a single day’s food intake may misestimate OOHF consumption.

Previous work has also explored demographic trends in the OOHF sector, showing a higher OOHF contribution primarily to those who are male [21], younger [5, 6, 21, 22], come from more deprived backgrounds [5, 22–24], have a higher Body Mass Index (BMI) [22] and those who have children [22, 25]. However, evidence also suggests demographic differences in the pattern of OOHF sources, with individuals from a high socioeconomic status (SES) spending more money on restaurants and cafés and those from a low SES spending more money on takeaways and fast food [25]. This suggests that previous studies, which have focused on location of food consumption only [2, 3, 7] and have therefore not captured takeaways consumed at home, may have misestimated SES trends for OOHF sector contributions to diet. Studies that have attempted to quantify the contribution that the OOHF makes to population diet are also limited by failing to recruit nationally representative samples [4, 5], basing estimation on a small number of diet recalls [4, 26] and not accounting for weekday vs. weekend variability in OOHF use [13] or seasonality [26].

In light of existing evidence, the contribution of the OOHF sector to individual total dietary intakes is unclear and may be misestimated in previous research. Information on detailed food intakes at the eating occasion-level over multiple days, linked to OOH nutrient composition and location of purchasing, would allow us to understand better the energy and nutrient intakes in the OOHF sector.

Therefore, the aim of the present study was to quantify the absolute and relative contribution that OOHF sources make to average daily nutrient intake and diet quality and the extent to which this differs across key demographics for the general adult population of England.

## Methods

### Study design and sample

Study protocols were pre-registered and described in detail on the Open Science Framework (https://osf.io/5n84w). Minor protocol deviations are described in Supplementary Information S1. Ethical approval was granted by the University of Bristol Faculty of Social Sciences and Law Research Ethics Committee (Ref.14629). In brief, we purposely sampled adults residing in England registered on the Prolific online research platform (prolific.com), stratifying the sample to ensure sufficient sub-group sizes for comparisons by age, sex and education.

We conducted a multi-stage online survey between September 2023 and May 2024. Participants self-reported demographics at JISC Online Surveys (onlinesurveys.ac.uk) and were invited to complete five, 24-hour dietary recalls via Prolific over the following 2-3 weeks, with sample boosts for non-response over a proceeding 2-3 weeks. Participants received ∼£0.60 per questionnaire and ∼£3.50 per dietary recall completed. The target sample size for recruitment was determined using a simulation approach detailed in the pre-registered protocol (https://osf.io/5n84w). Simulations were based on mean (standard deviation, SD) eating occasion size (kcal/occasion) and frequencies of eating out data from years 2008-2019 of the UK NDNS [7]. A total sample of N=1000 with five dietary recalls and reporting four eating occasions per day (with a conservative estimate of 0.5 OOH eating occasions a week), provides precision to detect at least a 60-kcal difference in energy intakes between OOH and non-OOH eating occasions using 95% confidence intervals (CIs). Simulation models assumed full participation, so we oversampled to account for non-response and attrition (≤20% [27, 28]) and aimed to recruit N=1200. We completed four waves of data collection, covering three seasons, with the demographic questionnaire, dietary recall and sample boost invites constituting one wave.

### Demographic data

Demographic questionnaires were designed using questions from established national food surveys, such as the Food Standards Agency’s Food and You survey [29], the Office for National Statistics Living Costs and Food Survey [30] and the UK NDNS [31, 32]. Participants reported their current age in years [31, 32], which was categorised as 18-29 / 30-44 / 45-59 / 60+ years; sex as male, female or prefer not to say, which was categorised as male / female [33] (prefer not to say categorised as male / female based on pre-screening participant demographic data provided by Prolific (n=4)); ethnicity as White, Black, Asian, Mixed, or Other [34], which was categorised as White / Black, Asian, Mixed, or Other combined; general health status as very good / good / fair / bad / very bad [31, 32]; current marital status, which was categorised as married, cohabiting or in civil partnership / single, widowed or divorced [29]; children in the household, which was categorised as yes / no [29]. Body mass index (BMI) was calculated from self-reported weight in kilograms (kg) and height in metres (m) as kg/m^2^ [31, 32] and categorised as <25 / 25-29.9 / 30+ kg/m^2^.

Socioeconomic status was measured by self-reported total household income, highest attained level of education (below UK A-level/UK A-level equivalent and above), current or last occupation and index of multiple deprivation (IMD). Equivalised household income was calculated to adjust for household composition and size and categorised as <£19,000 / £19,000-£31,999 / £32,000-£63,999 / £64,000-£95,999 / >£96,000 [35]. Occupation was categorised using the simplified National Statistics Socio-Economic Classification (NS-SEC) as higher / intermediate / routine/manual / not classifiable [36]. Economic activity status [37] was categorised as active (in or looking for employment) / inactive (student or unemployed and not looking for employment). Participants reported their lower layer super output area (LSOA) name [38], which was used to assign a 2011 rural or urban classification [39] and 2019 English IMD quintile (a measure of geographical area deprivation [40]). Dietary recall day was categorised as weekday (Monday-Friday) / weekend day (Saturday and Sunday); and time of year was categorised as meteorological seasons autumn (September 1^st^ – November 30^th^) / winter (December 1^st^ – February 28^th^) / spring (March 1^st^ – May 31^st^) [41].

### Dietary data

Dietary intakes were self-reported using Intake24, an online, open source 24hr recall tool used for the UK NDNS [27]. Participants recalled from the previous day (midnight to midnight) what, when, where from, and how much they ate and drank in meals or snacks (hereafter: eating occasions). In line with best practice, five non-consecutive recalls covering weekdays and weekends [27, 42, 43], were requested over a 2-3 week period, following the demographic questionnaire. Recall invitations across the entire study were aimed at representing all days of the week equally. Non-response to a recall invite prompted a re-invite during a subsequent 2-3 week ‘sample boost’ phase. Participants also reported frequency with which they typically eat or drink OOH in the demographics questionnaire [29].

#### Defining OOH food and drink

Out-of-home food and drink was defined in line with UK government legislation: “*foods and drinks (excluding alcohol without accompanying food) that can be purchased outside the home, either physically or digitally, that are ready for immediate consumption and that can be consumed in outlet premises, at home or elsewhere”* [44]. We excluded pre-packaged ready-to-eat items (e.g. pre-packaged sandwiches and crisps), to be consistent with how the UK government have defined out-of-home food in recent legislation [44].

Intake24 was modified to request eating occasion-level data on the category and name of outlet from which the majority of food and drink in an eating occasion came from, and whether the majority of this came ‘ready-to-eat’; defined for the participant as, “…*items from outside the home that came already cooked or prepared, with no further cooking, heating or preparation required before being eaten or drank…*”. Outlet category options (Supplementary Table S1) were defined by research team consensus drawing on previous studies exploring OOHF outlet use within the UK [25, 45–50]. Outlet categories selected by participants were cross-checked with the outlet name provided and recategorized, if necessary, by researchers.

To ensure eating occasions reported as ‘ready-to-eat’ fit the definition of eating occasions containing OOHF (i.e. excluding pre-packaged items), all eating occasions reported as ready-to-eat were manually checked by researchers and recoded OOHF or non-OOHF, as appropriate. Eating occasions reported as ready-to-eat from supermarkets, local/convenience stores and non-food shops, were assigned as non-OOH unless the participant also reported consuming the food/drink at the outlet location (assumed to be at the outlet café or restaurant).

#### Updating nutrient data

In Intake24, participants select a food and drink item that most closely matches items with nutritional composition data in the UK Nutrient Databank [27]. When no match can be found, participants provide a text description of the ‘missing’ food/drink, noting brand and portion sizes. Missing food items (n=750) were manually updated by researchers based on product nutritional information identified online [51]. Portion sizes were based on participant reported values or by using average portion sizes [52, 53]. The UK Nutrient Databank does not include up-to-date or complete OOH outlet-specific nutritional information on all OOHF products sold in the UK, therefore, nutritional information was supplemented using data from MenuTracker (version: September 2022) [54]. MenuTracker is a database containing nutritional information of menu items served by large (over 250 employees) UK OOHF chains. All foods in OOH eating occasions that had nutritional data available in MenuTracker were manually updated (n=1192/2837 (42%) OOH eating occasions), by one researcher (JG) and checked by another (NT). For outlets in MenuTracker with missing items consumed by participants, nutritional information was obtained from the outlet website. Further details on the use of MenuTracker for updating OOH nutritional data can be found in Supplementary Table S1.

#### Dietary data quality checking

In line with previous use of Intake24 [27, 55], daily energy intakes between 400-4000kcal were considered plausible energy intakes and any days outside these were removed (as such cut-offs are arbitrary, we applied an additional 10% buffer, removing: <360kcal (n=32 (0.6%)); >4400 kcal (n=70 (1.2%))). Dietary energy misreporting was also assessed via an individualised method [56] using a ratio of reported average daily energy intake to estimated energy requirement, calculated from standard equations [57] (see analysis protocol for full details). Additionally, portion sizes, total fat, saturated fat, sugar, and salt intakes (in grams) of individual items, excluding items updated using MenuTracker, were compared against maximum reported food group intakes in NDNS years 2008-2019 [7]. Dietary recalls containing foods with portion sizes and nutrient intakes exceeding these maximums (plus 10%) were removed (n=30 (0.5%)).

Dietary data quality in non-OOH eating occasions were also checked for food selection errors. Approximately 10% of all non-OOH eating occasions (n=1771) were randomly selected, examined for errors (e.g. ‘pea soup’ was initially typed in but ‘peas, boiled’ was chosen from the Intake24 food list), and manually amended with more appropriate food items [51]. Portion sizes were based on participant reported values or by using average portion sizes [52, 53]. Eating occasion energy intakes were compared between the original and corrected versions, indicating no significant differences (range: 0-3% energy) (Supplementary Table S2) and therefore the remaining unexamined eating occasions were considered acceptable [58].

## Statistical analysis

### Descriptives

All statistical analyses were performed using Stata version 18 (*StataCorp LLC, College Station, TX, USA*). Categorical variables were described as n(%) and continuous variables with the use of mean (standard error (SE)) if approximately normally distributed (based on histogram) or median (interquartile range (IQR): quartile 1, quartile 3) otherwise. To represent the wider population in England, all sample results were population-weighted by age, sex, ethnicity and highest education level, using England 2021 Census data [59] (Supplementary Table S3). Any day containing one or more OOH eating occasions was termed an ‘OOH day’.

Using dietary data, frequency of OOH eating occasions or days and their contribution to population energy intakes per week (by weighting weekday and weekend contributions and projecting to a 7-day week) were reported for the overall sample and within each demographic group. For allowing comparisons with existing data, weekly energy intake contributions from OOHF were also calculated but restricted to only those who reported OOHF sector use. Differences in energy intake (primary outcome) between OOH and non-OOH eating occasions or days within the overall sample and by key demographics (age, sex, BMI, and socioeconomic indicators) were tabulated. Energy intake was the primary outcome as it is a key factor in bodyweight regulation [60] and has direct relevance to public health obesity policy [61]. Secondary outcomes, including differences in nutrient intakes (total fat, saturated fat, total sugars, salt) between OOH and non-OOH eating occasions or days, were tabulated for descriptive purposes only. Average energy and nutrient intakes were also tabulated by outlet type for all OOH eating occasions, to quantify contribution from different OOHF sources.

To assess daily intakes compared with nutrient requirements on OOH and non-OOH days, the percentage of participants exceeding UK guidelines was calculated for females = 70g total fat; 20g saturated fat; males = 90g total fat; 30g saturated fat [62]; both sexes = 90g total sugars [63]; and 6g salt according to UK guidelines [64] and 5g salt according to WHO guidelines [65].

### Multi-level modelling

To test our main hypothesis, a series of random-intercept multilevel models were run to test descriptive differences in energy intakes between OOH and non-OOH 1) eating occasions (models 1.0-1.8) and 2) days (models 2.0-2.8). All models were run using maximum likelihood estimation [66], meaning participants who contributed fewer than five dietary recalls (n=380) were retained for analyses under a Missing At Random (MAR) assumption. Multilevel models were run using Stata’s *mixed* command.

For eating occasions, we initially tested whether a three-level model (eating occasions at level-1, nested within days at level-2, nested within participants at level-3) or a two-level model (eating occasions at level-1, nested within participants at level-2) should be used. Based on comparison of AIC and BIC values and model deviance, a two-level model was a better fit (Supplementary

Information S2). The initial 2-level model specified energy intake (kcal/occasion) as the outcome variable with a random-intercept for each person but no exposure variables. An intraclass correlation coefficient (ICC) was reported to show between-and within-person variation in eating occasion energy intakes. To test whether differences in OOH vs. non-OOH eating occasion energy intakes were potentially confounded, we first ran a random-intercept multilevel model in a sample restricted to those with complete covariate data. Energy intake was the outcome, and eating occasion type (OOH or non-OOH) was the only exposure variable (model 1.0). Model 1.1 additionally included all potential confounders as covariates at level-1 (time of year/season and week/weekend day) and level-2 (age, sex, ethnicity, BMI category, having a child at home, education, IMD, urban/rural location, NS-SEC and income). Model covariates were chosen based on literature linking demographic characteristics with differences in OOHF intakes [8, 25, 35, 67–71]. The effect estimates were compared to assess for evidence of confounding (models 1.0-1.1). To quantify how much energy intake variation is explained by the fixed effects (demographic covariates) and by both fixed and random effects (clustering within persons), marginal and conditional R^2^ were calculated respectively for model 1.1.

To test whether energy differences in OOH vs. non-OOH eating occasions varied by certain demographic groups, we further adjusted model 1.1 with interaction variables between eating occasion type (OOH/non-OOH) and age (model 1.2); sex (model 1.3); BMI category (model 1.4); education (model 1.5), IMD (model 1.6); NS-SEC (model 1.7); or income (model 1.8). Interaction term coefficients were the primary estimates of interest in models 1.2-1.8. Graphical model diagnostics were performed for each model to assess assumptions of level-1 and level-2 residual normality and level-1 residual homoscedasticity.

For day-level analysis, models 2.0-2.8 replicated models 1.0-1.8 but replaced eating occasion-level energy intakes with day-level energy intakes, eating occasion type as day type (OOH/non-OOH) and were additionally adjusted for frequency per day of OOH eating occasions to account for people eating OOHF more than once on the same day.

#### Sensitivity analysis

To explore sample selection bias, we report differences in the average number of eating occasions and days of diet submitted by demographic groups. We also checked for differences in average energy intakes between participants completing one, two, three, four or five recalls, complete cases (all five days of diet), a randomly selected four days of diet, a random three days of diet, a random two days of diet and a random one day of diet, in line with current NDNS methods [27]. Frequency of misreporting status was tabulated to observe for differences by demographic. To explore whether model associations related to potential energy intake misreporting, models 1.1-1.8 and day-level models 2.1-2.8 were repeated including misreporting status (under-reporting/plausible/over-reporting).

#### Accounting for multiple testing

The strength of evidence for associations was assessed by interpreting the size of the β and precision of their 95% confidence interval [72]. However, to account for multiple testing, the Benjamini-Hochberg procedure was applied with a false discovery rate (FDR) of 0.05 [73]. In this method, p-values for each key estimate from models 1.0-1.8 and 2.0-2.8 (and associated sensitivity analysis models), were ranked from smallest to largest. Each individual p-value was then compared to its Benjamini-Hochberg critical value, calculated for each p-value separately as (p-value rank/number of tests)*FDR i.e. (p-value rank/42)*0.05. To aid interpretation, we report both raw p-values and adjusted p-values based on the calculated Benjamini-Hochberg critical values (Supplementary Tables S10 and S12). The adjusted p-values were calculated as raw p-value*(number of tests/p-value rank). Adjusted p-values <0.05 were considered to reflect evidence of statistical significance.

## Results

### Sample characteristics

A total of N=1436 participants agreed to take part in the study. Complete demographic data were available for n=1403 (98%), of which n=1232 (86%) provided at least one dietary recall and were therefore included in multilevel analysis. From those, unweighted sample characteristics showed n=624 (51%) were female, n=1106 (90%) were of white ethnicity and were generally balanced across age groups and area deprivation scores (Table 1). Most participants reported having a BMI >25kg/m^2^ (58%; with 25% with BMI≥30 kg/m ^2^)^1^, having further or degree level education (51%), an equivalised household income above £19,000 (61%) and an intermediate or routine/manual occupation (50%).

**Table 1.** Unweighted characteristics of participants included in Models 1.0-1.8 and 2.0-2.8 (n=1232).

| Demographic | n (%) |
| --- | --- |
| **Sex** |  |
| Female | 624 (51%) |
| Male | 608 (49%) |

<sup>1</sup> A small number of participants (n=36) reported an underweight BMI (<18.5 kg/m<sup>2</sup>) and were therefore grouped under BMI <25kg/m<sup>2</sup>.

|  |  |
| --- | --- |
| **Ethnicity** |  |
| White | 1106 (90%) |
| Asian | 58 (5%) |
| Black | 28 (2%) |
| Mixed | 33 (3%) |
| Other | 7 (1%) |
| **Age (years) <sup>a</sup>** |  |
| 18-29 | 302 (25%) |
| 30-44 | 306 (25%) |
| 45-59 | 307 (25%) |
| 60+ | 317 (26%) |
| **BMI (kg/m<sup>2</sup>) <sup>b</sup>** |  |
| <25 | 521 (42%) |
| 25-29.9 | 401 (33%) |
| ≥30 | 310 (25%) |
| **Highest education** |  |
| No or compulsory school-level qualifications (basic) | 600 (49%) |
| University entry qualifications and vocational equivalents (further) | 130 (11%) |
| Degree level qualifications (degree) | 502 (41%) |
| **Equivalised total household income (£) <sup>c</sup>** |  |
| <19,000 | 481 (39%) |
| 19,000-31,999 | 376 (31%) |
| 32,000-63,999 | 327 (27%) |
| 64,000-95,999 | 39 (3%) |
| ≥96,000 | 9 (1%) |
| **Simplified NS-SEC analytic class** |  |
| 1 (Higher managerial, administrative and professional) | 535 (43%) |
| 2 (intermediate) | 307 (25%) |
| 3 (routine/manual) | 305 (25%) |
| Not classifiable <sup>d</sup> | 85 (7%) |
| **IMD score** |  |
| Quintile 1 (most deprived) | 224 (18%) |
| Quintile 2 | 272 (22%) |
| Quintile 3 | 249 (20%) |
| Quintile 4 | 261 (21%) |
| Quintile 5 (least deprived) | 226 (18%) |
| **Has child (<18 years) living at home** | 382 (31%) |
| **Living in urban area** | 1012 (82%) |
| **Marital status** |  |
| Married, cohabiting or in civil partnership | 789 (64%) |
| Single, widowed or divorced | 443 (36%) |
| **Economically active (in or seeking employment)** | 906 (74%) |
| **Self-reported general health status** |  |
| Very good | 179 (15%) |
| Good | 604 (49%) |
| Fair | 384 (31%) |
| Bad | 61 (5%) |
| Very bad | 4 (<1%) |
| **Energy misreporting status** |  |
| under-reporters | 791 (64%) |
| plausible reporters | 386 (31%) |
| over-reporters | 55 (4%) |

BMI – Body Mass Index; NS-SEC – National Statistics Socioeconomic Category; IMD – Index of Multiple Deprivation. a – ‘Not classifiable’ due to never having worked/full-time student/occupation not stated/occupation not classifiable for other reasons. Percentages may not add up to 100% due to rounding. Population weighted median (IQR) values for age: 47 (32, 59) years; BMI: 26 (23.2, 29.7) kg/m<sup>2</sup>; equivalised household income: £24,667 (£16,000, £38,000).

### Dietary data

In the overall sample, on average, participants completed a total of 4.5 (SE 0.03) dietary recalls (69% participants completing all 5 dietary recalls) (Supplementary Tables S4). Average daily energy intakes were unaffected by differences in the total number of dietary recalls completed (Supplementary Table S5). More dietary recalls were completed by participants of White vs. Asian, Black, Mixed or Other ethnicities (mean 4.5 (SE 0.03) vs 4.0 (0.13) days) and older vs. younger participants (60+ years: 4.6 (0.05) vs 18-29 years: 4.2 (0.08)) (Supplementary Table S6). Most dietary intakes were recorded on weekdays (64%) versus weekends (36%), and during winter (36%) or spring (35%) vs. autumn (29%).

Participants reported a total of 4.5 (SE 0.04) eating occasions per day (Supplementary Table S7) and 2.2 (IQR: 0, 5) or 2.5 (1, 5) OOH eating occasions per week based on recalls or the questionnaire respectively. Descriptively, eating out was more frequent in women versus men (median 2.5 vs 2.0 occasions per week), those with Asian, Black, Mixed or Other ethnicity versus White ethnicity (3.3 vs 2.1 occasions per week), younger age groups (18-29: 2.8 vs 45+: 2.0 occasions per week), higher BMI categories (≥30 kg/m^2^ : 2.4 vs <25 kg/m^2^ : 2.0 occasions per week), higher equivalised household incomes (>£96,000: 4.5 vs £64,000-£95,999: 2.7 vs <£19,000: 2.0 occasions per week) and higher education levels (degree level: 2.5 vs basic: 2.0 occasions per week) (Supplementary Table S7).

### Energy and nutrient intakes

In the overall sample, without adjusting for potential confounders, average energy intakes during OOH eating occasions were higher compared to non-OOH eating occasions (median 440kcal vs 294kcal, Supplementary Table S7). Similarly, OOH eating occasions compared to non-OOH eating occasions were descriptively higher in absolute intakes of total fat (median 17g vs 9g), saturated fat (5g vs 3g), total sugar (14g vs 11g) and salt (1.1g vs 0.5g) (Figure 1 and Supplementary Table S7). The same pattern was observed at day-level, with OOH days versus non-OOH days being higher in energy (1774 vs 1575 kcal), total fat (68g vs 58g), saturated fat (24g vs 21g), total sugars (72g vs 63g) and salt (4.8g vs 4.2g) (Figure 1 and Supplementary Table S6). OOHF contributed a median of 11.0 (IQR: 0, 23.4) % energy to total weekly energy intakes (weighted by weekday/weekend days) overall and 16.9 (9.6, 30.2) % energy in those who reported ≥1 OOH eating occasion. Higher percentages of participants reported exceeding recommended daily intakes of fat, saturated fat, total sugars and salt on OOH days compared with non-OOH days (>70g fat (female): 43% vs 29%, >90g fat (male): 29% vs 16%; >20g saturated fat (female): 62% vs 53%, >30g saturated fat (male): 36% vs 28%; >90g total sugars: 33% vs 26%; >6g salt: 32% vs 18%; > 5g salt: 51% vs. 35%).

**Figure 1.**
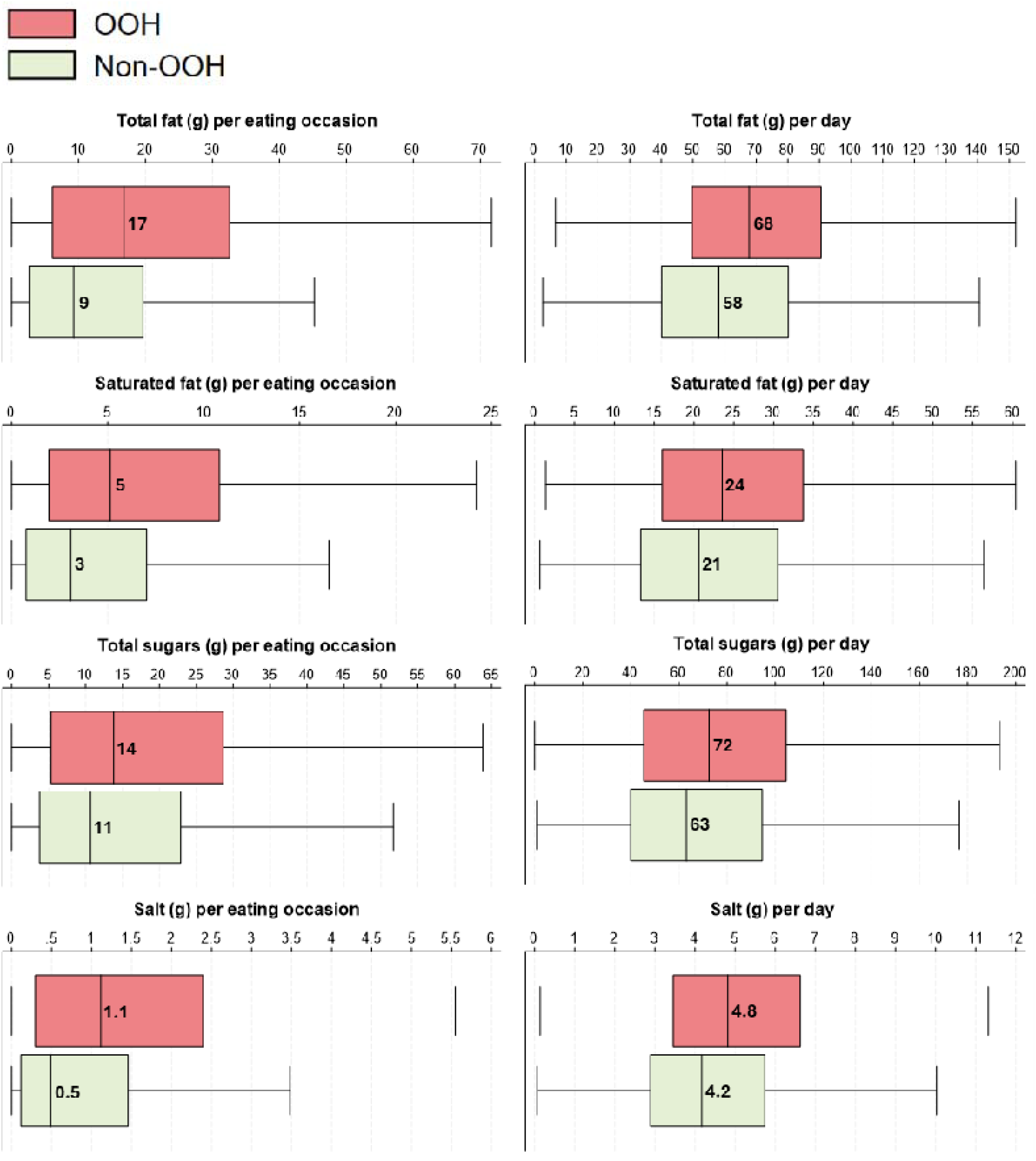
Population-weighted nutrient intake boxplots for participants included in eating occasion-level models 1.0-1.8 and day-level models 2.0-2.8. Median intakes are indicated in each box. OOH – out-of-home; Non-OOH – non-out-of-home

In the overall sample, median total fat intakes as a percentage of total energy intake (%TE) were higher OOH at both eating occasion-level (34%TE vs 30%TE) and day-level (34%TE vs 33%TE), saturated fat intakes (%TE) were slightly higher OOH at the eating occasion-level (11%TE vs 10%TE) but the same at day-level (12%TE vs 12%TE), and total sugar intakes (%TE) were lower OOH at the eating occasion-level (13%TE vs 17%TE) but the same at day-level (16%TE vs 16%TE) (Supplementary Tables S8 and S9).

Median energy intakes at both OOH and non-OOH eating occasions were observed to be higher in younger age groups versus older (OOH: 18-29 years 488 kcal vs 60+ years 377kcal; non-OOH: 18-29 years 489 kcal vs 60+ years 264kcal; Supplementary Table S7). No other differences in energy and nutrient intakes were observed across other demographics (Supplementary Table S7). Median energy and nutrient intakes at the day-level were also consistently higher on OOH days but there was no clear trend in energy intake by age at the eating-occasion level (Supplementary Table S6).

### Out-of-home outlet-specific nutrient intakes

The most commonly reported outlet types for OOH eating occasions were fast food/takeaway/street food outlets (25%), supermarkets (17%), café/coffee house/tea rooms (11%), restaurants (10%) and bakery/sandwich shops (10%; Figure 2). As well as being most common, eating occasions at fast food/takeaway/street food outlets were also highest in energy (median: 828 kcal; contributing 47% total daily energy on days this outlet type was visited), followed by restaurants which provided 771 kcal (49% total daily energy) and pubs/bars/clubs providing 589 kcal (38% total daily energy; Table 2). Fast food/takeaway/street food outlets also provided the highest amounts of total fat, saturated fat and salt intakes (median intakes per eating occasion: 35g, 9g and 2.3g respectively) amongst all outlet types (Table 2).

**Table 2.**
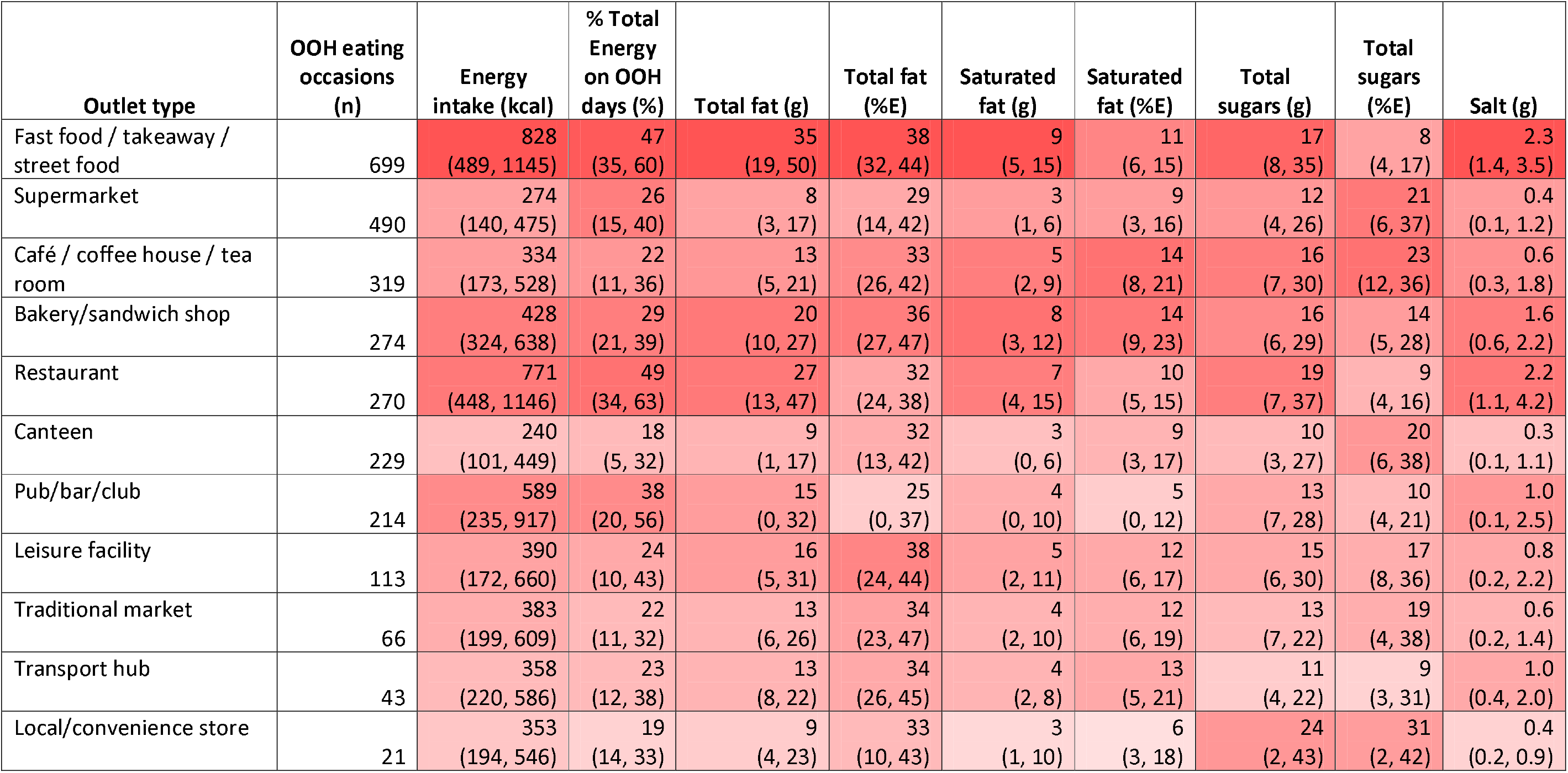
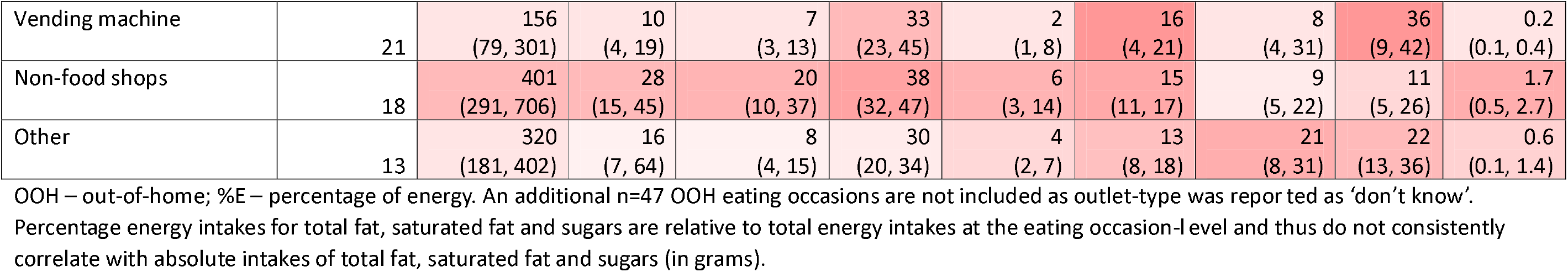
Outlet-type frequency of use (n) and median (IQR) energy and nutrient intakes by outlet type for days that include OOH eating occasions (n=2,837) for participants included in models 1.0-1.8 and 2.0-2.8 (N=1232). Colour coding: Each column value was ranked from 1 to 14 (based on increasing values). Ranks for energy/nutrient intake columns were multiplied by the rank of eating occasion frequency to indicate the overall contribution of each outlet type to OOH energy and nutrient intakes. Cells are shaded darker (linearly) for higher combined values and lighter for lower combined values, with darker shades thus indicating the greatest combined contribution to OOH energy and nutrient intakes. Data are presented in order of descending reporting frequency of each outlet type.

| Outlet type | OOH eating occasions (n) | Energy intake (kcal) | % Total Energy on OOH days (%) | Total fat (g) | Total fat (%E) | Saturated fat (g) | Saturated fat (%E) | Total sugars (g) | Total sugars (%E) | Salt (g) |
| --- | --- | --- | --- | --- | --- | --- | --- | --- | --- | --- |
| Fast food / takeaway / street food | 699 | 828<br>(489, 1145) | 47<br>(35, 60) | 35<br>(19, 50) | 38<br>(32, 44) | 9<br>(5, 15) | 11<br>(6, 15) | 17<br>(8, 35) | 8<br>(4, 17) | 2.3<br>(1.4, 3.5) |
| Supermarket | 490 | 274<br>(140, 475) | 26<br>(15, 40) | 8<br>(3, 17) | 29<br>(14, 42) | 3<br>(1, 6) | 9<br>(3, 16) | 12<br>(4, 26) | 21<br>(6, 37) | 0.4<br>(0.1, 1.2) |
| Café / coffee house / tea room | 319 | 334<br>(173, 528) | 22<br>(11, 36) | 13<br>(5, 21) | 33<br>(26, 42) | 5<br>(2, 9) | 14<br>(8, 21) | 16<br>(7, 30) | 23<br>(12, 36) | 0.6<br>(0.3, 1.8) |
| Bakery/sandwich shop | 274 | 428<br>(324, 638) | 29<br>(21, 39) | 20<br>(10, 27) | 36<br>(27, 47) | 8<br>(3, 12) | 14<br>(9, 23) | 16<br>(6, 29) | 14<br>(5, 28) | 1.6<br>(0.6, 2.2) |
| Restaurant | 270 | 771<br>(448, 1146) | 49<br>(34, 63) | 27<br>(13, 47) | 32<br>(24, 38) | 7<br>(4, 15) | 10<br>(5, 15) | 19<br>(7, 37) | 9<br>(4, 16) | 2.2<br>(1.1, 4.2) |
| Canteen | 229 | 240<br>(101, 449) | 18<br>(5, 32) | 9<br>(1, 17) | 32<br>(13, 42) | 3<br>(0, 6) | 9<br>(3, 17) | 10<br>(3, 27) | 20<br>(6, 38) | 0.3<br>(0.1, 1.1) |
| Pub/bar/club | 214 | 589<br>(235, 917) | 38<br>(20, 56) | 15<br>(0, 32) | 25<br>(0, 37) | 4<br>(0, 10) | 5<br>(0, 12) | 13<br>(7, 28) | 10<br>(4, 21) | 1.0<br>(0.1, 2.5) |
| Leisure facility | 113 | 390<br>(172, 660) | 24<br>(10, 43) | 16<br>(5, 31) | 38<br>(24, 44) | 5<br>(2, 11) | 12<br>(6, 17) | 15<br>(6, 30) | 17<br>(8, 36) | 0.8<br>(0.2, 2.2) |
| Traditional market | 66 | 383<br>(199, 609) | 22<br>(11, 32) | 13<br>(6, 26) | 34<br>(23, 47) | 4<br>(2, 10) | 12<br>(6, 19) | 13<br>(7, 22) | 19<br>(4, 38) | 0.6<br>(0.2, 1.4) |
| Transport hub | 43 | 358<br>(220, 586) | 23<br>(12, 38) | 13<br>(8, 22) | 34<br>(26, 45) | 4<br>(2, 8) | 13<br>(5, 21) | 11<br>(4, 22) | 9<br>(3, 31) | 1.0<br>(0.4, 2.0) |
| Local/convenience store | 21 | 353<br>(194, 546) | 19<br>(14, 33) | 9<br>(4, 23) | 33<br>(10, 43) | 3<br>(1, 10) | 6<br>(3, 18) | 24<br>(2, 43) | 31<br>(2, 42) | 0.4<br>(0.2, 0.9) |
| Vending machine | 21 | 156<br>(79, 301) | 10<br>(4, 19) | 7<br>(3, 13) | 33<br>(23, 45) | 2<br>(1, 8) | 16<br>(4, 21) | 8<br>(4, 31) | 36<br>(9, 42) | 0.2<br>(0.1, 0.4) |
| Non-food shops | 18 | 401<br>(291, 706) | 28<br>(15, 45) | 20<br>(10, 37) | 38<br>(32, 47) | 6<br>(3, 14) | 15<br>(11, 17) | 9<br>(5, 22) | 11<br>(5, 26) | 1.7<br>(0.5, 2.7) |
| Other | 13 | 320<br>(181, 402) | 16<br>(7, 64) | 8<br>(4, 15) | 30<br>(20, 34) | 4<br>(2, 7) | 13<br>(8, 18) | 21<br>(8, 31) | 22<br>(13, 36) | 0.6<br>(0.1, 1.4) |

OOH – out-of-home; %E – percentage of energy. An additional n=47 OOH eating occasions are not included as outlet-type was reported as ‘don’t know’.
Percentage energy intakes for total fat, saturated fat and sugars are relative to total energy intakes at the eating occasion-level and thus do not consistently correlate with absolute intakes of total fat, saturated fat and sugars (in grams).

**Figure 2.**
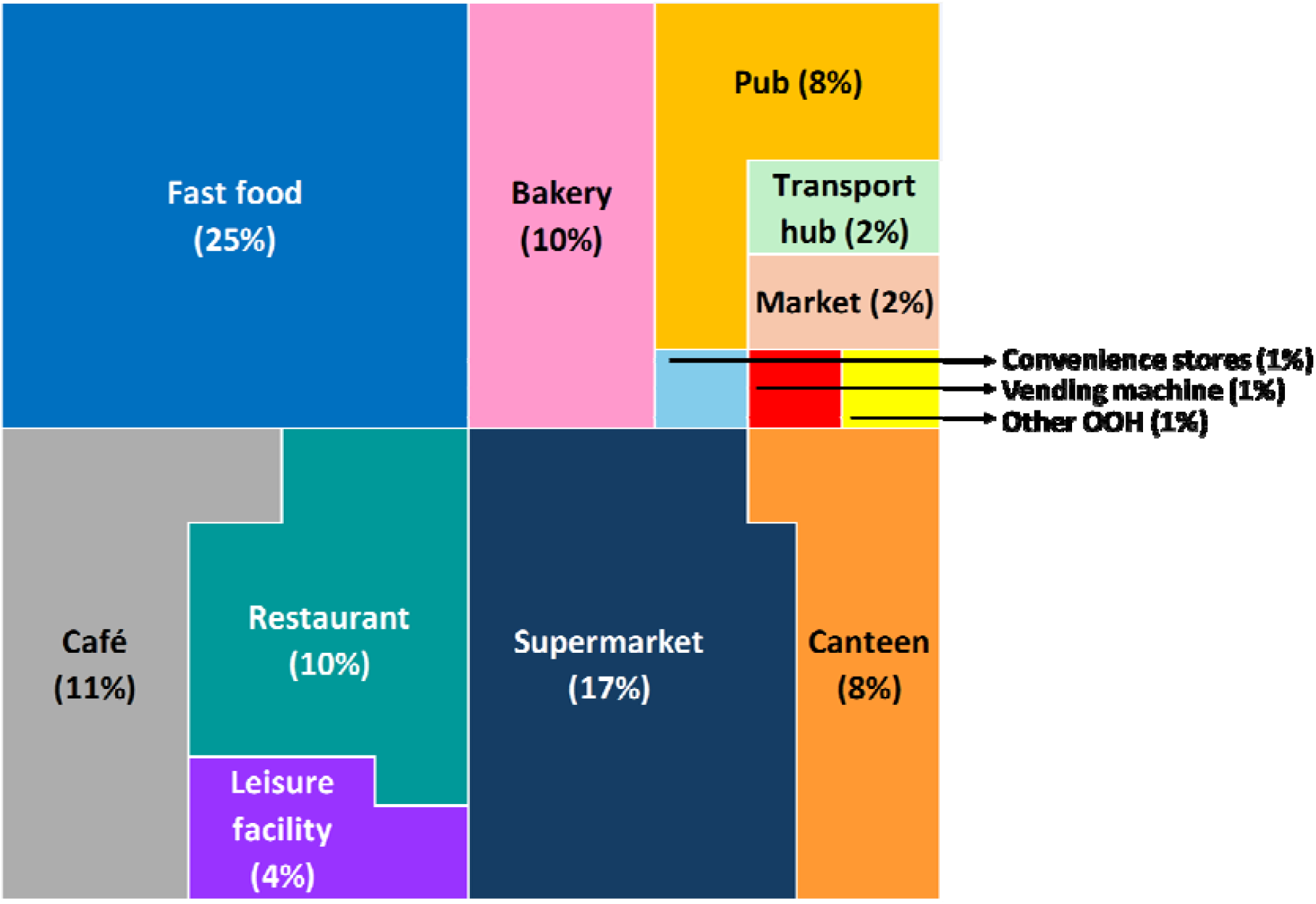
Proportions of out-of-home (OOH) eating occasions by outlet type. OOH – out-of-home

### Multilevel modelling

#### Eating occasion-level (Models 1.0-1.8)

Most variation in energy intake at eating occasions was within-person before (91.3%) and after adjusting for covariates (93.9%). Demographic variables explained only 6.0% of the variance and combined with individual-level differences (random intercepts), they explained 11.8% of the variance. OOH eating occasions were found to be β=199 [95%CI: 174, 224] kcal (p<0.001) and 196 [171, 221] kcal (p<0.001) higher in energy than non-OOH eating occasions prior to, and after, full covariate adjustment respectively (Model 1.0-1.1; Figure 3 and Supplementary Table S10). Strong evidence of associations remained after accounting for multiple testing. No evidence was found indicating that differences in OOH and non-OOH eating occasions varied by demographic (age, sex, BMI, education, IMD, occupational social class and equivalised household income) (Models 1.2-1.8, Supplementary Table S10).

**Figure 3.**
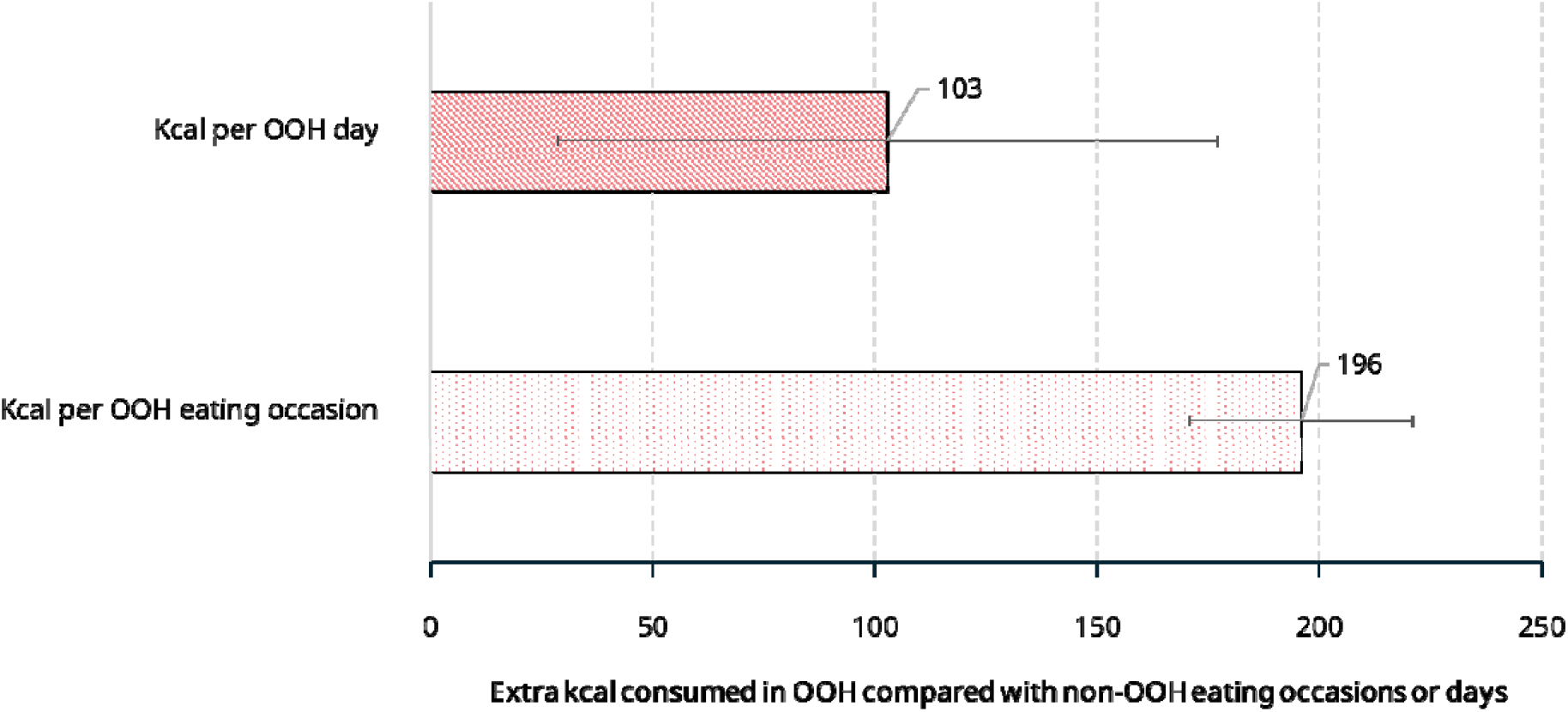
Additional energy (kcal) consumed in out-of-home (OOH) compared with non-OOH eating occasions and days (Models 1.1 and 2.1). Confounder adjusted estimates and 95% CIs are shown. OOH – out-of-home

#### Day-level (Models 2.0-2.8)

Most variation in daily energy intake was within-persons before (57.8%) and after adjusting for covariates (60.5%). Demographic variables explained 8.2% of the variance in daily energy intake and combined with individual-level differences (random intercepts) explained 44.5%. Days containing OOH foods were found to be β=111 [95%CI: 38, 183] kcal (p=0.003) and 103 [29, 177] kcal (p=0.006) higher in energy than days without OOHF prior to, and after, covariate adjustment respectively (Model 2.0-2.1; Figure 3 and Supplementary Table S10). However, after adjusting for covariates and accounting for multiple testing, day-level differences in energy intakes between OOH and non-OOH days were no longer statistically significant at p<0.05 (adjusted p=0.063). No evidence was found that indicated the size of differences in energy intakes between OOH and non-OOH days varied by demographic characteristics (Models 2.2-2.8; Supplementary Table S10).

#### Sensitivity Analysis

Average daily under-reporting and over-reporting of energy intake was estimated to occur in 64% and 4% participants respectively (Supplementary Table S11). Energy under-reporting was more common in younger age groups (18-29 years: 70% vs 60+ years: 59%), males (72% vs females 57%), participants with higher BMI (≥30 kg/m^2^ : 74% vs <25: 55%), and lower IMD quintile (IMD 1 (most deprived): 71% vs IMD 5 (least deprived): 62%) (Supplementary Table S11). However, findings from Models 1.1-1.8 and 2.1-2.8 remained the same after further adjustment for misreporting status and after accounting for multiple testing (Supplementary Table S12).

## Discussion

This study provides the first quantification of the contribution of the OOHF sector to the dietary intakes of adults in England using eating occasion-level data. Across more than 24,000 eating occasions and 5,400 person-days of diet, the findings reveal that OOHF consumption is associated with higher energy and nutrient intakes compared to non-OOHF consumption. Specifically, OOH eating occasions contributed 196 kcal more energy than non-OOH occasions and contained more total fat, saturated fat, sugar, and salt. Fast food and restaurant outlets emerged as the primary contributors to OOHF consumption. While partial compensation for higher OOHF intake occurred across other non-OOH meals in a day, the net effect was still an increase in daily energy intake on OOHF days of 103 kcal after adjusting for confounding factors (although this difference did not remain significant after correction for multiple testing). These patterns were consistent across demographic groups, underscoring the broad impact of the OOHF sector on dietary quality.

In the present study, we estimated the average (median) contribution of the OOHF sector to daily energy intake as being between 11.0% (total sample) and 16.9% (including OOHF consumers only). Previous studies vary in their average estimate of the contribution that the OOHF sector makes to daily energy intake, ranging from 12-50% [5, 6, 21, 26, 74]. Our estimation is similar to a government report of a representative sample of 2,146 UK adults (12%) between 2019-2023, although our analysis benefited by enhancing the energy and nutritional information of foods obtained from the OOHF sector. Differences in estimated contribution between other studies is likely to be due to socio-cultural differences between countries sampled and methods used. For example, a recent unpublished report estimated that the OOHF sector contributed an average of 300kcal per day per person (15% of energy intake for a 2000kcal daily intake) among UK adults [5]. However, the approach used sampled regular OOH users only, examined purchase data (as opposed to consumption) and included packaged sandwiches bought from supermarkets as an OOHF product, all of which will increase the estimated contribution of the OOHF sector to national diet. Other methodological differences which may explain why our study findings differ to others include reporting the median rather than mean, which was more appropriate to use due to the very skewed distribution of OOH eating occasions. Mean OOHF contributions to total weekly energy intakes were 15.7% compared to the reported median of 11.0%. In addition, differences in estimates may be due to our decision to weight estimated contribution by weekday vs. weekend (as frequency of eating out differs between the two) and seasonality [16]. Weighting data by weekday vs. weekend provides a more accurate estimate of the OOHF sector contribution, as unweighted data may reflect variation in data collection rather than true variation in OOHF versus non-OOHF energy intakes. Previous studies have also not consistently accounted for seasonal variation in OOHF sector use [26].

Our findings are consistent with international and UK-based research indicating that OOHF is associated with excess energy consumption and poorer nutrient profiles [5, 8, 14, 25, 49]. A higher frequency of OOHF consumption by younger and more affluent individuals also aligns with results from previous studies [21]. The majority of OOH eating occasions in the current study (25%) were from fast food settings (both chain and independent) and were high in energy (47% of total daily energy on OOH days), which aligns with previous work in the UK [5, 6]. However, our study advances prior research by using detailed, outlet-specific nutrient data and a robust multilevel modelling approach, which gives more confidence in the associations, as they are not confounded by measured person level characteristics, which were held constant in multilevel models. Our findings also showed that supermarkets, coffee shops, bakeries and restaurants were frequent sources of OOH eating occasions, but it was eating occasions in restaurants that were the second highest in energy intake after fast food outlets. This is unlike previous research showing that supermarkets were the second highest OOH contributor to energy intake [5], although that analysis used purchase data and included pre-packaged sandwiches suggesting these may be important methodological choices in the description of OOH food.

Our findings reinforce the need for strengthened policy interventions targeting the OOHF sector. That OOHF occasions are energy-dense across all demographics indicates that population-wide, rather than subgroup-specific, regulatory interventions are warranted. Designing policies that can specifically target the OOHF sector to induce reformulation and subsequently reduce energy density should therefore be pursued [75]. Mandatory menu calorie labelling [76] (which is already implemented in England for large chains [44]); portion control measures [77]; banning price and volume promotions of HFSS products in the OOHF sector [78, 79]; reducing the price of healthier menu items [80]; and taxing products sold in the OOH sector [81], could be effective policies worth exploring, particularly for fast food and restaurant outlets, which were commonly visited and provided high energy, fat and salt foods. Healthcare providers and public health campaigns should incorporate guidance on reducing high-energy OOHF consumption.

However, focusing only on calories or energy density alone will be insufficient to address the poor-quality food consumed from OOHF sources, as our findings also highlight that eating occasions from the OOHF sector are high in fat, sugar and salt. Thus, improvement in the nutritional quality, as well as the amount of food offered in these settings is important. Currently, there is no mechanism for assessing the nutritional quality of an eating occasion. The UK Nutrient Profile Model [82] is designed to assess the quality of individual foods, while OOH eating occasions represent combinations of foods. Future research could explore quantifying the nutritional quality alongside the amount of energy in OOH eating occasions.

For researchers, our study highlights the importance of detailed food source identification and updated and expanded nutrient databases in national dietary surveys to reflect variation in portion size and nutritional composition of foods in the OOHF sector. Surveillance systems should adopt similar methodologies to monitor the evolving food environment effectively. For example, this could include expanding the current proposal under the Food Data Transparency Partnership, which requires large food retailers in the UK to disclose the ratio of healthy to unhealthy products sold, to also include OOHF businesses [83]. Future studies should extend data collection to include summer months to comprehensively capture seasonal variations in OOHF consumption. The development of scalable methodologies for real-time dietary tracking and food source identification, such as Ecological Momentary Assessment (EMA) [84], would also be informative. Finally, our classification of OOHF followed existing UK legislation, which excludes pre-packaged ready-to-eat items such as sandwiches and crisps [44]. Food to go from supermarkets is one of the major contributors to OOHF purchasing [5]. Therefore, future research should quantify the contribution of pre-packaged ready-to-eat foods currently excluded, as well as the contribution of both meals and non-meals (i.e., snacks) to OOH and non-OOH occasions to better understand their impact on overall energy and nutrient intake.

A major strength of this study is that we recruited a stratified sample reflective of England’s adult population (without restricting it only to OOHF consumers), by applying weights by key demographics. We collected an average of 4.5 days of dietary recall per participant, exceeding many national nutrition surveys in recall depth, and analyses accounted for within-and between-person variation. Additionally, the study captured seasonal variation and stratified intake patterns by detailed demographic factors, offering a more nuanced understanding of OOHF consumption. These methods allowed for a clearer differentiation of the nutritional impact of OOHF across various outlet types and provide a replicable framework for future dietary surveillance. We employed Intake24 enhanced by the MenuTracker database to provide more accurate and up-to-date nutritional profiling for foods obtained from the OOHF sector. The integration of MenuTracker addressed a critical limitation of existing datasets, which often rely on generic food composition tables [7].Additionally, the multilevel modelling framework accounted for within- and between-person variation in intake which allowed us to explore the independent effect of the OOHF sector on energy intake independent of person-level characteristics.

However, several limitations must also be acknowledged. The reliance on an online panel (Prolific) may limit generalisability due to self-selection bias and the exclusion of individuals without access to the internet or the required technology. In addition, registrants who had not provided information to Prolific on age, sex and highest education level attained, were automatically excluded from the eligible sample pool when stratifying by these variables during recruitment. Samples obtained using Prolific cannot be fully representative of adults in England, with no ability to implement random sampling methods as study adverts within the platform can only be targeted to specific groups or demographics (quota sampling) and not randomly selected individuals. Data collection did not include summer months, potentially underestimating annual OOHF consumption [16]. We quantified misreporting of energy intake, which showed under-reporting of energy intake, particularly among younger, male, and higher-BMI participants, although sensitivity analyses suggested that this impacted minimally on the overall findings (i.e., relative contributions to diet). Although under-reporting of energy intake would result in a lower estimated energy intake during both OOH and non-OOH occasions, the absolute difference between the two we observed is likely to be underestimated. Although we used MenuTracker to manually update nutritional information for eating occasions assigned as OOH, this was possible for only 42% of OOH eating occasions, as these contained food obtained from chain restaurants included in MenuTracker, and no non-OOHF data. Future research should aim to specifically assess energy and nutritional quality from foods sold in independent food businesses, which is currently missing.

## Conclusions

The OOHF sector contributes substantially to the dietary energy and nutrient intake of adults in England. Our study, using detailed eating occasion-level data, where we held person-level confounding constant, showed that OOH eating occasions are 196 kcal higher in energy compared to non-OOH eating occasions. Our findings also indicated that OOH eating occasions contain higher fat, saturated fat, sugar and salt than non-OOH eating occasions. These patterns were consistent across demographic groups and highlight the widespread dietary impact of the OOHF sector. Findings support the need for strengthened, population-wide policy interventions, particularly targeting fast food and restaurant outlets, to improve dietary quality and reduce diet-related disease.

## Data Availability

Currently all data produced in the present study are available upon reasonable request to the authors.

## Footnotes

1 A small number of participants (n=36) reported an underweight BMI (<18.5 kg/m^2^) and were therefore grouped under BMI <25kg/m^2^.

