## Supplementary material for "The contribution of the out-of-home food (OOHF) sector to the national diet: a cross-sectional survey with repeated 24-hour recalls of adults in England (2023-2024)"

**Supplementary Materials**

**Supplementary Information**

**Supplementary Information S1**. Deviations from methods or analysis protocols (https://osf.io/5n84w/).

1) If differences exist between the methods and analysis protocols (for example, the defining of research questions and the definition of OOH food/drinks within this study), the later-published analysis protocol takes precedence.
2) We were unable to update nutrient data during OOH eating occasions using the most up-to-date version of the MenuTracker (MT) database [1]. This was due to data sharing policy reviews taking place at the University of Cambridge (MT proprietor) coinciding with study data collection and analysis timelines. We instead used the September 2022 version of the MT database for updating OOH nutrient data that was available from conducting previous research.
3) We were unable to access a version of the FoodDB database [2] for updating non-OOH foods/drinks due to data sharing policy reviews at the University of Oxford (FoodDB proprietor) coinciding with study data collection and analysis timelines.
4) For the purposes of presenting data more coherently within the manuscript, eating occasion- and day-level main analysis model labels have been reversed relative to numbering detailed in the protocol, such that eating occasion-level models are now labelled Models 1.1-1.8 and day-level models now labelled Models 2.1-2.8.
5) To observe effects of covariate adjustment on effect estimates, Models 1.0 and 2.0 have been added to demonstrate differences in energy between out-of-home and non-out-of-home eating occasions and days in those with complete covariate data, prior to the adjustment with covariates in Models 1.1 and 2.1 respectively.
6) We have additionally presented marginal and conditional R^2^ values for models 1.1 and 2.1 to demonstrate the energy intake variance explained by model fixed effects (demographic covariates) and model fixed effects plus random effects (demographic covariates plus random intercepts).

**Supplementary Information S2**. Eating occasion-level model fit.

To determine whether eating occasion-level energy intakes were most appropriately modelled using a three-level model (eating occasions at level-1, nested within days at level-2, nested within participants at level-3) or a two-level model (eating occasions at level-1, nested within participants at level-2), we ran a three-level and a two-level empty-means random-intercept model [42] with eating occasion-level energy intake as the outcome and with no predictors. We then compared model-derived values for Aikake Information Criterion (AIC), Bayesian Information Criterion (BIC), and model deviance (-2×log-likelihood; -2LL). The empty means two-level random intercept model with eating occasions at level-1 nested within persons at level-2 was found to be the most parsimonious model (-2LL: 4914; AIC: 4920; BIC: 4936) compared to the empty means three-level random intercept model with eating occasions at level-1, nested within days at level-2, nested within persons at level-3 (-2LL: 4914; AIC: 4922; BIC: 4942). Eating occasion models 1.0-1.8 were thus based on a two-level rather than a three-level random intercept model.

**Supplementary Tables**

**Supplementary Table S1**. Study variables, their categories and additional information on data cleaning and/or calculation.

| **Variable/Data** | **Categories** | **Additional information** |
| --- | --- | --- |
| Sex | 1) Female;  2) Male |  |
| Ethnicity | 1) White;  2) Asian, Black, Mixed or Other | Asian, Black, Mixed and other ethnicities merged into single group due to small sample size. |
| Age (years) | 1) 18-29;  2) 30-44;  3) 45-59;  4) 60+ |  |
| Bodyweight (kg) | N/A | Implausible bodyweight values defined as bodyweight <30 kg or >400kg [3]. |
| Height (m) | N/A | Implausible height values defined as height <1.2m or >2.2m [3]. |
| BMI (kg/m^2^) | 1) <25 (normal weight);  2) 25-29.9 (overweight);  3) 30+ (obesity) | After removing implausible bodyweight and height data, Body mass index (BMI) was calculated as: bodyweight in kilograms/(height in metres)^2^ |
| Minutes of MVPA per day | N/A | Weekly time spent in moderate-to-vigorous physical activity >21 hours and <35 hours were truncated to 21 hours, and values >35 hours categorised as implausible [4]. |
| Equivalised total household income (£) | 1) <19,000;  2) 19,000-31,999;  3) 32,000-63,999;  4) 64,000-95,999;  5) >96,000 [5]. | Equivalised total household income is a measure of income adjusted for household composition and size. Calculated as total household income divided by the sum of the modified Organisation for Economic Co-operation and Development (OECD) equivalence values for each household member (First adult: 1.0; Additional adult: 0.5; Child aged 14 and over: 0.5; Child aged 0-13: 0.3 [6]). |
| IMD quintile | 1) quintile 1 (most deprived);  2) quintile 2;  3) quintile 3;  4) quintile 4;  5) quintile 5 (least deprived) | Lower Layer Super Output Area (LSOA) names were used to assign a 2019 English Index of Multiple Deprivation (IMD) quintile. IMD is the official measure of relative deprivation for small areas in England [7]. |
| Self-reported general health status | 1) Very good;  2) good;  3) fair;  4) bad;  5) very bad |  |
| Marital status | 1) Married, cohabiting or in civil partnership;  2) Single, widowed or divorced |  |
| Has child <18 years living at home | 1) Yes;  2) No | Classified as ‘yes’ if participants reported both: 1) yes to having children and 2) reporting a number of children (<18 years) living in the household greater than zero. This avoided miscategorising participants (for the purposes of our analysis) as having children when those children were over 18 years old. |
| Highest education | 1) No or compulsory school-level qualifications (basic);  2) University entry qualifications and vocational equivalents (further);  3) Degree level qualifications (degree) [8, 9]. |  |
| Energy misreporting status | 1) under-reporter;  2) plausible-reporter;  3) over-reporter. | See analysis protocol for full derivation details (https://osf.io/5n84w/). |
| Urban/rural location | 1) Urban;  2) Rural | LSOA names were used to assign a 2011 rural or urban geographical classification [10]. |
| Economic activity level | 1) Active (in or looking for employment);  2) Inactive (student or unemployed and not looking for employment) | Economic activity status (classifying whether the participant contributes to the UK labour market [11]) was defined as ‘active’ if the participant reported being employed or unemployed but actively looking for work, and ‘inactive’ otherwise. Economically inactive groups typically include students, people looking after family and home, long-term or temporarily sick and disabled, retired people and discouraged workers [11]. |
| Occupational social class: Simplified NS-SEC | 1) Higher managerial, administrative and professional;  2) Intermediate;  3) Routine and manual.  4) Not classifiable | Standard Occupational Classification 2010 (SOC2010) codes were used to assign a three-class simplified National Statistics Socio-Economic Classification (NS-SEC) [12]. NS-SEC is the official socio-economic classification index used in the UK constructed to measure employment relations and conditions of different occupations. Intermediate occupations include clerical, sales, service and technical occupations that do not involve general planning or supervisory powers [12]. A fourth residual category (‘not classifiable’) was designated for those reporting never having worked, full-time students, ‘occupations not stated’, or ‘occupations not classifiable for other reasons’ [12]. |
| Time of week | 1) weekday (Monday-Friday);  2) weekend day (Saturday and Sunday) |  |
| Season/Time of year | 1) Autumn;  2) Spring;  3) Winter | Defined as meteorological autumn (September 1^st^ – November 30^th^), winter (December 1^st^ – February 28^th^), and spring (March 1^st^ – May 31^st^) |
| Out-of-home outlet type | 1) Supermarket;  2) Local/convenience store (e.g. Premier, Spar, Nisa, etc.);  3) Traditional market/delicatessen/butchers/fishmongers/farm shop;  4) Bakery/sandwich shop (e.g. Greggs, Cooplands, Pret a Manger, etc.);  5) Transport hub (e.g. petrol station, motorway service station, airport, etc.);  6) Restaurant;  7) Café/coffee house/tea rooms;  8) Pub/bar/club;  9) Fast food/takeaway outlet/street food outlet;  10) Non-food shops (e.g. pharmacies, discount stores, off licences, etc.);  11) Leisure facility (e.g. tourist attraction, gym, cinema, concert venue, hotels, etc.) ;  12) Vending machine;  13) Canteen (e.g. at work, college, university, etc.);  14) Grown at home/allotment (includes tap water);  15) Other (e.g. online store, food bank, etc.);  16) Don’t know. | Outlet category response options were reached by research team consensus and based on previous studies exploring OOH outlet use within the UK [13–19]. |
| Energy and nutrient data (for out-of-home foods reported at outlets with nutrient data available on Menutracker 2022 database) | N/A | Portion sizes were taken from MenuTracker for items as they appeared on the menu and were adjusted proportionally by the ratio of the serving size versus amount consumed reported by the participant. In cases where MenuTracker contained nutritional information for only single food units (e.g. per chicken wing) but with no corresponding portion size, average portion sizes were assumed [20, 21]. Participant data were then updated using the total consumed portion size they reported; except where they explicitly specified the number of single units consumed. In cases where nutritional information per 100g was available on MenuTracker for identical menu items with varying portion size labels, i.e. small/regular/large, participant data were updated using the portion size (in grams) reported as consumed. If nutritional information per 100g was not available on MenuTracker for identical menu items with varying portion size labels, i.e. small/regular/large, regular portion size nutritional data were applied instead. |

**Supplementary Table S2**. Dietary data quality assessment results for non-out-of-home eating occasion energy intakes. Differences between raw and corrected energy did not exceed 10% in any of sex, ethnicity, age or education demographics.

|  |  | **Raw energy (kcal)** | | **Corrected energy (kcal)** | | **Difference  (corrected-raw energy; %)** | |
| --- | --- | --- | --- | --- | --- | --- | --- |
| **Demographic** | **n** | **mean** | **median** | **mean** | **median** | **mean** | **median** |
| **Overall** | 1771 | 371 | 287 | 369 | 286 | -1.0% | -0.2% |
| **Sex** |  |  |  |  |  |  |  |
| Female | 919 | 337 | 252 | 337 | 254 | 0.0% | 0.9% |
| Male | 852 | 408 | 341 | 403 | 339 | -1.1% | -0.8% |
| **Ethnicity** |  |  |  |  |  |  |  |
| White | 1646 | 373 | 288 | 371 | 287 | -0.5% | -0.3% |
| Asian, Black, Mixed and Other | 125 | 350 | 263 | 348 | 263 | -0.6% | 0.0% |
| **Age (years)** |  |  |  |  |  |  |  |
| 18-29 | 328 | 421 | 326 | 421 | 329 | 0.0% | 0.9% |
| 30-44 | 386 | 343 | 282 | 346 | 285 | 0.9% | 1.1% |
| 45-59 | 441 | 403 | 284 | 401 | 276 | -0.5% | -2.8% |
| 60+ | 616 | 339 | 281 | 333 | 281 | -1.8% | 0.0% |
| **Highest education level** |  |  |  |  |  |  |  |
| No or compulsory school-level qualifications (basic) | 864 | 399 | 288 | 395 | 285 | -1.0% | -1.0% |
| University entry qualifications and vocational equivalents (further) | 182 | 328 | 288 | 327 | 289 | -0.3% | 0.3% |
| Degree level qualifications (degree) | 725 | 349 | 286 | 348 | 287 | -0.3% | 0.3% |

**Supplementary Table S3**. Population demographic proportions for age, sex, ethnicity and highest education level for adjusting sample results to represent the larger population.

Data calculated from the England 2021 Census [22]. Values presented do not add up to 100% due to rounding. Ethnicity has been categorised into ‘White’ and ‘Asian, Black, Mixed and Other’ due to individual Asian, Black, Mixed and Other ethnic sub-group weightings being inappropriately small for use with a total recruited sample of N=1232.

|  | **White ethnicity** | | | | **Asian, Black, Mixed and Other ethnicity** | | | |
| --- | --- | --- | --- | --- | --- | --- | --- | --- |
|  | **Female** | | **Male** | | **Female** | | **Male** | |
| **Age (years)** | **Below A-level education** | **A-level and above education** | **Below A-level education** | **A-level and above education** | **Below A-level education** | **A-level and above education** | **Below A-level education** | **A-level and above education** |
| **18-29** | 2.3% | 4.9% | 2.9% | 4.3% | 0.6% | 1.7% | 0.7% | 1.4% |
| **30-44** | 3.2% | 6.8% | 3.8% | 5.8% | 1.1% | 2.0% | 1.0% | 1.7% |
| **45-59** | 5.1% | 5.7% | 5.2% | 5.3% | 1.0% | 1.1% | 1.0% | 1.0% |
| **60+** | 10.6% | 4.4% | 7.9% | 5.2% | 0.9% | 0.4% | 0.7% | 0.5% |

**Supplementary Table S4**. Total number of 24-hour dietary recalls returned by participants included in Models 1.0-1.8 and 2.0-2.8.

| **Total number of 24-hour dietary recalls ^a^** | **n (%)** |
| --- | --- |
| 1 | 46 (4%) |
| 2 | 56 (5%) |
| 3 | 88 (7%) |
| 4 | 190 (15%) |
| 5 | 852 (69%) |

a - maximum of five possible dietary recalls

**Supplementary Table S5**. Median (Q1, Q3) daily energy intake (kcal) calculated from different numbers of completed dietary recalls per participant (all those included in models 1.0-1.8 and 2.0-2.8 (including one, two, three, four or five days of diet), complete cases (five days of diet), four randomly selected days of diet, three randomly selected days of diet, two randomly selected days of diet and one randomly selected days of diet) to explore potential for selection bias for participants included in Models 1.0-1.8 and 2.0-2.8.

|  | **Number of completed 24-hr dietary recalls** | | | | | |
| --- | --- | --- | --- | --- | --- | --- |
|  | **1-5 days** | **5 complete days** | **4 random days** | **3 random days** | **2 random days** | **1 random day** |
| n | 1232 | 852 | 1042 | 1130 | 1186 | 1232 |
| Daily energy intake (kcal) | 1658 (1324, 2007) | 1664 (1339, 1990) | 1672 (1332, 2032) | 1659 (1327, 2027) | 1651 (1290, 2053) | 1635 (1212, 2058) |

**Supplementary Table S6**. Population weighted median (Q1, Q3) energy and nutrient intakes from OOH days (n=1,897) and non-OOH days (n=3,545) by demographic characteristics for participants included in Models 1.0-1.8 and 2.0-2.8 (N=1,232).

|  | **Mean (SE) number of 24-hr dietary recalls** | **Median (IQR) number of OOH days**  **per week^a^** | **Daily energy intake (kcal)** | | **Total fat (g)** | | **Total saturated fat (g)** | | **Total sugars (g)** | | **Total salt (g)** | |
| --- | --- | --- | --- | --- | --- | --- | --- | --- | --- | --- | --- | --- |
|  |  |  | **OOH** | **non-OOH** | **OOH** | **non-OOH** | **OOH** | **non-OOH** | **OOH** | **non-OOH** | **OOH** | **non-OOH** |
| **All** | 4.5 (0.03) | 2.0  (0, 3.7) | 1774  (1390, 2262) | 1575  (1188, 1989) | 68  (50, 91) | 58  (40, 80) | 24  (16, 34) | 21  (13, 31) | 72  (45, 105) | 63  (40, 95) | 4.8  (3.4, 6.6) | 4.2  (2.9, 5.7) |
| **Sex** |  |  |  |  |  |  |  |  |  |  |  |  |
| Female | 4.5 (0.04) | 2.0  (0.7, 3.7) | 1659  (1276, 2067) | 1462  (1120, 1834) | 64  (46, 84) | 55  (38, 75) | 23  (16, 32) | 20  (13, 29) | 69  (44, 99) | 61  (39, 91) | 4.5  (3.2, 6.2) | 3.9  (2.7, 5.4) |
| Male | 4.4 (0.04) | 1.7  (0.0, 3.7) | 1923  (1533, 2459) | 1680  (1265, 2137) | 73  (54, 98) | 62  (42, 85) | 25  (17, 36) | 22  (14, 32) | 76  (47, 111) | 65  (40, 97) | 5.2  (3.8, 7.1) | 4.5  (3.0, 6.2) |
| **Ethnicity** |  |  |  |  |  |  |  |  |  |  |  |  |
| White | 4.5 (0.03) | 2.0  (0.0, 3.7) | 1772  (1393, 2260) | 1582  (1189, 1991) | 68  (50, 91) | 59  (40, 80) | 24  (16, 34) | 21  (13, 31) | 73  (45, 105) | 63  (40, 95) | 4.9  (3.5, 6.6) | 4.2  (2.9, 5.8) |
| Asian, Black, Mixed and Other | 4.0 (0.13) | 2.5  (0.7, 4.3) | 1793  (1262, 2319) | 1422  (1051, 1862) | 68  (48, 93) | 53  (37, 75) | 21  (15, 32) | 17  (12, 25) | 66  (45, 106) | 54  (32, 82) | 4.2  (3.2, 6.4) | 3.3  (2.1, 4.8) |
| **Age (years)** |  |  |  |  |  |  |  |  |  |  |  |  |
| 18-29 | 4.2 (0.08) | 2.5  (0.0, 4.5) | 1777  (1438, 2305) | 1584  (1194, 1997) | 68  (50, 96) | 60  (41, 81) | 24  (17, 33) | 21  (13, 31) | 77  (48, 108) | 65  (43, 99) | 4.9  (3.5, 6.9) | 4.2  (2.8, 5.7) |
| 30-44 | 4.3 (0.07) | 2.0  (1.0, 3.7) | 1784  (1368, 2285) | 1611  (1213, 2046) | 68  (51, 93) | 59  (43, 83) | 23  (16, 35) | 21  (14, 31) | 68  (41, 100) | 64  (39, 100) | 4.9  (3.4, 6.9) | 4.1  (2.9, 5.6) |
| 45-59 | 4.6 (0.05) | 1.7  (0.0, 3.3) | 1835  (1415, 2317) | 1579  (1163, 2001) | 68  (50, 88) | 59  (40, 80) | 24  (16, 34) | 21  (13, 31) | 75  (50, 112) | 60  (38, 94) | 5.1  (3.6, 6.6) | 4.3  (2.9, 5.9) |
| 60+ | 4.6 (0.05) | 2.0  (0.0, 3.7) | 1721  (1318, 2140) | 1481  (1132, 1835) | 65  (47, 87) | 55  (38, 77) | 23  (16, 33) | 20  (13, 29) | 70  (43, 101) | 63  (41, 89) | 4.3  (3.1, 6.0) | 4.1  (2.8, 5.6) |
| **BMI (kg/m^2^)** |  |  |  |  |  |  |  |  |  |  |  |  |
| <25 | 4.5 (0.05) | 2.0  (0.0, 3.7) | 1765  (1409, 2218) | 1596  (1215, 1999) | 68  (50, 93) | 61  (42, 82) | 23  (16, 34) | 21  (13, 31) | 74  (48, 106) | 69  (44, 102) | 4.7  (3.3, 6.4) | 4.1  (2.8, 5.6) |
| 25-29.9 | 4.4 (0.06) | 2.0  (0.0, 3.8) | 1831  (1390, 2343) | 1550  (1143, 1947) | 67  (51, 89) | 55  (38, 78) | 24  (16, 34) | 20  (13, 29) | 74  (44, 109) | 60  (37, 89) | 5.0  (3.5, 6.7) | 4.2  (2.9, 5.8) |
| ≥30 | 4.5 (0.06) | 2.0  (0.7, 3.7) | 1736  (1342, 2229) | 1568  (1182, 1977) | 68  (49, 89) | 59  (41, 81) | 24  (15, 33) | 21  (14, 32) | 67  (42, 100) | 58  (35, 87) | 5.0  (3.6, 6.8) | 4.3  (3.0, 6.0) |
| **Marital status** |  |  |  |  |  |  |  |  |  |  |  |  |
| Married, cohabiting or in civil partnership | 4.5 (0.04) | 2.0  (0.0, 3.7) | 1763  (1376, 2245) | 1571  (1199, 1983) | 67  (49, 89) | 57  (41, 79) | 23  (16, 34) | 21  (14, 30) | 72  (44, 105) | 63  (40, 93) | 4.9  (3.5, 6.6) | 4.2  (3.0, 5.7) |
| Single, widowed or divorced | 4.5 (0.05) | 2.0  (0.0, 3.8) | 1819  (1404, 2309) | 1587  (1162, 1998) | 68  (50, 96) | 60  (38, 84) | 24  (16, 34) | 21  (13, 31) | 74  (49, 105) | 63  (40, 99) | 4.8  (3.4, 6.6) | 4.1  (2.7, 5.8) |
| **Child status** |  |  |  |  |  |  |  |  |  |  |  |  |
| Has child/children living at home | 4.5  (1.0) | 2.0  (0.7, 3.7) | 1847  (1439, 2401) | 1583  (1225, 2022) | 71  (53, 95) | 59  (43, 81) | 25  (17, 36) | 22  (14, 31) | 72  (44, 108) | 64  (41, 94) | 5.2  (3.7, 7.0) | 4.3  (3.0, 5.8) |
| No child/children living at home | 4.4  (1.1) | 2.0  (0.0, 3.7) | 1747  (1360, 2207) | 1570  (1163, 1962) | 66  (48, 89) | 57  (39, 80) | 23  (16, 33) | 20  (13, 30) | 73  (46, 103) | 63  (39, 95) | 4.7  (3.3, 6.5) | 4.1  (2.8, 5.7) |
| **Equivalised household income (£)** |  |  |  |  |  |  |  |  |  |  |  |  |
| <19,000 | 4.5 (0.05) | 1.7  (0.0, 3.7) | 1772  (1396, 2319) | 1579  (1182, 2003) | 69  (50, 93) | 60  (39, 83) | 24  (16, 35) | 21  (13, 31) | 75  (48, 111) | 64  (40, 94) | 4.6  (3.3, 6.3) | 4.1  (2.8, 5.8) |
| 19,000-31,999 | 4.5 (0.06) | 2.0  (0.0, 3.3) | 1772  (1368, 2254) | 1610  (1200, 1991) | 68  (50, 92) | 60  (42, 81) | 24  (16, 34) | 21  (15, 30) | 70  (42, 104) | 65  (39, 97) | 4.9  (3.5, 6.8) | 4.3  (3.0, 5.9) |
| 32,000-63,999 | 4.4 (0.06) | 2.0  (0.7, 3.8) | 1775  (1395, 2178) | 1527  (1156, 1943) | 65  (49, 87) | 55  (38, 78) | 22  (15, 33) | 19  (12, 30) | 74  (47, 101) | 62  (39, 94) | 4.9  (3.5, 6.6) | 4.1  (2.7, 5.6) |
| 64,000-95,999 | 4.4 (0.16) | 1.7  (1.0, 4.5) | 1908  (1500, 2630) | 1623  (1272, 2034) | 73  (57, 102) | 60  (40, 80) | 25  (20, 36) | 22  (14, 33) | 68  (44, 93) | 54  (43, 90) | 5.3  (3.9, 7.0) | 4.3  (3.1, 5.7) |
| >96,000 | 4.1 (0.39) | 3.3  (2.7, 3.8) | 1435  (1188, 1737) | 1502  (1378, 1721) | 55  (45, 64) | 53  (50, 67) | 19  (19, 24) | 20  (15, 27) | 48  (35, 56) | 43  (30, 57) | 4.3  (3.3, 5.9) | 4.4  (3.0, 4.9) |
| **Highest education level** |  |  |  |  |  |  |  |  |  |  |  |  |
| No or compulsory school-level qualifications (basic) | 4.5 (0.04) | 1.7  (0.0, 3.7) | 1816  (1385, 2276) | 1547  (1158, 1969) | 70  (49, 94) | 56  (39, 78) | 24  (16, 36) | 20  (13, 30) | 74  (43, 112) | 61  (38, 90) | 4.8  (3.3, 6.8) | 4.1  (2.9, 5.8) |
| University entry qualifications and vocational equivalents (further) | 4.4 (0.04) | 2.0  (0.0, 3.7) | 1673  (1283, 2200) | 1553  (1169, 1965) | 63  (47, 86) | 56  (38, 79) | 23  (15, 33) | 20  (12, 29) | 67  (35, 99) | 62  (41, 89) | 4.8  (3.3, 6.4) | 4.2  (2.9, 5.8) |
| Degree level qualifications (degree). | 4.5 (0.05) | 2.0  (0.0, 3.7) | 1788  (1403, 2262) | 1596  (1200, 2001) | 67  (51, 88) | 60  (42, 82) | 24  (16, 33) | 21  (14, 31) | 73  (47, 101) | 65  (40, 99) | 4.9  (3.5, 6.7) | 4.2  (2.9, 5.7) |
| **IMD quintile** |  |  |  |  |  |  |  |  |  |  |  |  |
| 1 (most deprived) | 4.3 (0.09) | 1.7  (0.0, 3.3) | 1731  (1307, 2361) | 1533  (1151, 1977) | 66  (50, 94) | 57  (38, 80) | 24  (16, 35) | 20  (12, 31) | 68  (44, 102) | 60  (35, 92) | 4.9  (3.2, 6.7) | 4.2  (2.9, 5.7) |
| 2 | 4.5 (0.06) | 2.0  (1.0, 4.3) | 1849  (1385, 2309) | 1566  (1173, 1991) | 70  (48, 91) | 58  (39, 81) | 24  (16, 34) | 21  (13, 30) | 73  (44, 102) | 64  (42, 100) | 4.9  (3.6, 6.7) | 4.1  (2.8, 5.8) |
| 3 | 4.5 (0.07) | 2.0  (0.0, 3.7) | 1723  (1308, 2159) | 1630  (1223, 2043) | 64  (49, 87) | 61  (44, 84) | 23  (15, 33) | 22  (15, 33) | 75  (45, 108) | 65  (40, 98) | 4.8  (3.4, 6.5) | 4.3  (3.0, 6.0) |
| 4 | 4.6 (0.06) | 2.0  (0.0, 3.8) | 1784  (1455, 2242) | 1518  (1145, 1903) | 66  (50, 87) | 54  (38, 74) | 23  (16, 32) | 19  (12, 28) | 76  (48, 107) | 59  (37, 86) | 4.8  (3.5, 6.5) | 4.1  (2.8, 5.6) |
| 5 (least deprived) | 4.5 (0.07) | 2.0  (0.0, 3.3) | 1837  (1423, 2305) | 1628  (1219, 2001) | 69  (52, 92) | 60  (42, 81) | 23  (17, 34) | 22  (14, 31) | 69  (46, 104) | 67  (42, 99) | 4.9  (3.5, 6.8) | 4.1  (2.9, 5.6) |
| **Urban/rural location** |  |  |  |  |  |  |  |  |  |  |  |  |
| Urban | 4.5 (0.07) | 2.0  (0.0, 3.7) | 1772  (1395, 2257) | 1569  (1182, 1983) | 67  (50, 91) | 59  (40, 80) | 24  (16, 33) | 21  (13, 30) | 72  (45, 102) | 63  (40, 95) | 4.8  (3.5, 6.7) | 4.2  (2.9, 5.8) |
| Rural | 4.5 0.03) | 1.7  (0.0, 3.3) | 1779  (1362, 2297) | 1606  (1199, 1997) | 68  (48, 89) | 57  (39, 83) | 23  (16, 36) | 20  (13, 31) | 77  (44, 112) | 64  (39, 93) | 5.0  (3.4, 6.5) | 4.2  (2.8, 5.7) |
| **Economic activity** |  |  |  |  |  |  |  |  |  |  |  |  |
| Active | 4.4 (0.04) | 2.0  (0.0, 3.7) | 1793  (1402, 2305) | 1597  (1197, 2003) | 68  (50, 92) | 59  (41, 81) | 24  (16, 34) | 21  (13, 31) | 74  (46, 106) | 64  (40, 97) | 5.0  (3.5, 6.8) | 4.2  (2.9, 5.8) |
| Inactive | 4.6 (0.06) | 1.7  (0.0, 3.7) | 1728  (1308, 2142) | 1508  (1142, 1901) | 64  (48, 86) | 56  (38, 78) | 23  (16, 33) | 20  (13, 29) | 68  (41, 101) | 61  (39, 89) | 4.4  (3.0, 6.0) | 4.0  (2.8, 5.5) |
| **Simplified NS-SEC analytic class** |  |  |  |  |  |  |  |  |  |  |  |  |
| 1 (Higher managerial, administrative and professional) | 4.5 (0.04) | 2.0  (0.0, 3.7) | 1784  (1415, 2276) | 1601  (1200, 2001) | 69  (52, 91) | 59  (42, 80) | 24  (16, 34) | 21  (14, 30) | 72  (45, 102) | 62  (40, 96) | 5.0  (3.6, 6.7) | 4.2  (2.9, 5.7) |
| 2 (intermediate) | 4.5 (0.07) | 2.0  (0.0, 3.7) | 1767  (1351, 2231) | 1571  (1134, 1951) | 66  (47, 86) | 58  (39, 80) | 24  (16, 33) | 20  (13, 31) | 73  (44, 104) | 61  (38, 92) | 4.8  (3.3, 6.7) | 4.0  (2.9, 5.7) |
| 3 (routine or manual) | 4.4 (0.07) | 2.0  (0.0, 3.7) | 1801  (1374, 2305) | 1558  (1176, 2002) | 68  (48, 95) | 55  (37, 79) | 23  (15, 35) | 20  (13, 31) | 77  (51, 112) | 66  (40, 95) | 4.7  (3.3, 6.6) | 4.2  (3.0, 6.0) |
| Not classifiable | 4.4 (0.12) | 1.3  (0.0, 3.7) | 1599  (1290, 2007) | 1505  (1181, 1877) | 60  (45, 79) | 62  (40, 81) | 22  (16, 29) | 20  (13, 28) | 54  (33, 92) | 63  (41, 94) | 4.5  (3.0, 6.2) | 3.8  (2.7, 5.3) |

OOH – out-of-home; BMI – Body Mass Index; IMD – Index of Multiple Deprivation; NS-SEC – National Statistics Socio-Economic Classification based on occupation. a – weighted by weekday/weekend to account for potential differences in OOH use with time of week.

**Supplementary Table S7**. Population-weighted median (Q1, Q3) energy and nutrient intakes during OOH eating occasions (n=2,837) and non-OOH eating occasions (n=21,345) by demographic characteristics for participants included in Models 1.0-1.8 and 2.0-2.8 (N=1,232).

|  | **Mean (SE) eating occasions per day** | **Median (IQR) number of eating occasions**  **per week^a^** | | **Eating occasion energy (kcal)** | | **Total fat (g)** | | **Total saturated**  **fat (g)** | | **Total sugars (g)** | | **Total salt (g)** | |
| --- | --- | --- | --- | --- | --- | --- | --- | --- | --- | --- | --- | --- | --- |
|  |  | **OOH** | **non-OOH** | **OOH** | **non-OOH** | **OOH** | **non-OOH** | **OOH** | **non-OOH** | **OOH** | **non-OOH** | **OOH** | **non-OOH** |
| **All** | 4.5  (0.04) | 2.2  (0.0, 5.0) | 26.9  (21.0, 33.7) | 440  (215, 783) | 294  (129, 505) | 17  (6, 33) | 9  (3, 20) | 5  (2, 11) | 3  (1, 7) | 14  (5, 29) | 11  (4, 23) | 1.1  (0.3, 2.4) | 0.5  (0.1, 1.5) |
| **Sex** |  |  |  |  |  |  |  |  |  |  |  |  |  |
| Female | 4.4  (0.05) | 2.5  (0.7, 5.2) | 27.8  (21.8, 34.5) | 407  (209, 739) | 267  (118, 458) | 16  (7, 30) | 9  (2, 18) | 5  (2, 10) | 3  (1, 6) | 14  (5, 28) | 10  (4, 22) | 1.0  (0.3, 2.3) | 0.4  (0.1, 1.3) |
| Male | 4.6  (0.05) | 2.0  (0.0, 5.0) | 26.3  (20.3, 32.8) | 477  (230, 843) | 331  (149, 560) | 18  (6, 35) | 10  (3, 22) | 5  (2, 11) | 3  (1, 8) | 14  (6, 29) | 11  (4, 25) | 1.2  (0.3, 2.5) | 0.6  (0.1, 1.6) |
| **Ethnicity** |  |  |  |  |  |  |  |  |  |  |  |  |  |
| White | 4.5  (0.04) | 2.1  (0.0, 5.0) | 27.0  (21.0, 33.8) | 442  (215, 786) | 294  (128, 505) | 17  (6, 33) | 9  (3, 20) | 5  (2, 11) | 3  (1, 7) | 14  (5, 29) | 11  (4, 23) | 1.1  (0.3, 2.4) | 0.5  (0.1, 1.5) |
| Asian, Black, Mixed and Other | 4.1  (0.10) | 3.3  (0.5, 7.0) | 21.8  (15.4, 28.6) | 410  (217, 726) | 306  (149, 507) | 15  (7, 30) | 11  (3, 20) | 5  (2, 10) | 3  (1, 7) | 14  (5, 30) | 10  (4, 22) | 1.0  (0.3, 2.2) | 0.5  (0.1, 1.3) |
| **Age (years)** |  |  |  |  |  |  |  |  |  |  |  |  |  |
| 18-29 | 4.1  (0.07) | 2.8  (0.0, 6.2) | 23.3  (17.9, 30.0) | 488  (250, 856) | 489  (275, 813) | 19  (8, 35) | 11  (4, 22) | 6  (2, 12) | 4  (1, 8) | 17  (8, 32) | 12  (5, 26) | 1.2  (0.3, 2.7) | 0.6  (0.2, 1.6) |
| 30-44 | 4.4  (0.06) | 2.2  (0.9, 5.2) | 25.8  (20.1, 32.1) | 472  (238, 832) | 308  (151, 518) | 21  (8, 35) | 10  (4, 21) | 6  (2, 12) | 3  (1, 7) | 13  (4, 27) | 11  (4, 23) | 1.5  (0.3, 2.6) | 0.5  (0.1, 1.5) |
| 45-59 | 4.5  (0.08) | 2.0  (0.0, 4.7) | 28.1  (22.5, 34.6) | 438  (205, 779) | 284  (119, 501) | 15  (5, 32) | 9  (2, 19) | 5  (2, 10) | 3  (1, 7) | 13  (5, 30) | 11  (4, 23) | 1.0  (0.2, 2.3) | 0.5  (0.1, 1.5) |
| 60+ | 4.9  (0.06) | 2.0  (0.0, 4.6) | 30.3  (24.1, 35.9) | 377  (189, 683) | 264  (103, 457) | 13  (5, 27) | 8  (2, 18) | 4  (2, 9) | 3  (1, 6) | 13  (6, 25) | 9  (3, 21) | 0.8  (0.2, 2.0) | 0.4  (0.1, 1.3) |
| **BMI (kg/m^2^)** |  |  |  |  |  |  |  |  |  |  |  |  |  |
| <25 | 4.6  (0.05) | 2.0  (0.0, 5.0) | 27.5  (22.0, 34.0) | 427  (207, 771) | 294  (134, 503) | 15  (6, 32) | 9  (3, 20) | 5  (2, 10) | 3  (1, 7) | 14  (5, 29) | 12  (4, 24) | 0.9  (0.3, 2.3) | 0.5  (0.1, 1.4) |
| 25-29.9 | 4.5  (0.07) | 2.2  (0.0, 5.1) | 26.4  (20.6, 33.6) | 432  (214, 759) | 288  (120, 498) | 16  (6, 31) | 9  (2, 19) | 5  (2, 11) | 3  (1, 7) | 13  (6, 28) | 10  (3, 23) | 1.1  (0.3, 2.3) | 0.5  (0.1, 1.5) |
| ≥30 | 4.3  (0.07) | 2.4  (0.6, 5.0) | 26.7  (20.2, 32.9) | 483  (234, 824) | 306  (131, 518) | 20  (8, 35) | 10  (3, 21) | 6  (2, 12) | 3  (1, 8) | 15  (5, 29) | 10  (3, 23) | 1.4  (0.3, 2.6) | 0.5  (0.1, 1.6) |
| **Marital status** |  |  |  |  |  |  |  |  |  |  |  |  |  |
| Married, cohabiting or in civil partnership | 4.5  (0.04) | 2.2  (0.0, 5.0) | 27.2  (22.0, 33.8) | 427  (214, 766) | 290  (128, 498) | 17  (6, 32) | 9  (3, 19) | 5  (2, 11) | 3  (1, 7) | 13  (5, 28) | 11  (4, 23) | 1.1  (0.3, 2.4) | 0.5  (0.1, 1.5) |
| Single, widowed or divorced | 4.4  (0.07) | 2.1  (0.0, 5.1) | 26.0  (20.0, 33.5) | 464  (218, 828) | 305  (131, 521) | 17  (6, 33) | 10  (3, 21) | 5  (2, 11) | 3  (1, 7) | 15  (5, 30) | 11  (4, 24) | 1.2  (0.3, 2.4) | 0.5  (0.1, 1.5) |
| **Child status** |  |  |  |  |  |  |  |  |  |  |  |  |  |
| Has child/children living at home | 4.5  (0.04) | 2.0  (0.7, 5.0) | 27.1  (22.0, 33.8) | 477  (229, 844) | 302  (133, 507) | 20  (7, 38) | 10  (3, 20) | 6  (2, 12) | 3  (1, 7) | 16  (6, 31) | 11  (4, 23) | 1.4  (0.3, 2.7) | 0.5  (0.1, 1.5) |
| No child/children living at home | 4.5  (0.06) | 2.2  (0.0, 5.2) | 26.7  (20.8, 33.7) | 424  (212, 752) | 291  (126, 503) | 15  (6, 31) | 9  (3, 20) | 5  (2, 10) | 3  (1, 7) | 13  (5, 28) | 11  (4, 23) | 1.1  (0.3, 2.2) | 0.5  (0.1, 1.4) |
| **Equivalised household income (£)** |  |  |  |  |  |  |  |  |  |  |  |  |  |
| <19,000 | 4.5  (0.06) | 2.0  (0.0, 5.1) | 27.1  (20.6, 33.9) | 446  (245, 761) | 293  (120, 515) | 16  (7, 33) | 9  (2, 20) | 6  (2, 11) | 3  (1, 7) | 15  (6, 30) | 11  (4, 23) | 1.2  (0.3, 2.3) | 0.5  (0.1, 1.4) |
| 19,000-31,999 | 4.5  (0.06) | 2.0  (0.0, 4.7) | 27.1  (22.1, 34.0) | 471  (226, 822) | 297  (134, 513) | 18  (7, 35) | 9  (3, 20) | 6  (2, 11) | 3  (1, 7) | 14  (5, 30) | 11  (4, 23) | 1.2  (0.3, 2.5) | 0.5  (0.1, 1.5) |
| 32,000-63,999 | 4.5  (0.07) | 2.5  (1.0, 5.3) | 26.7  (20.1, 32.8) | 403  (195, 765) | 292  (134, 488) | 15  (5, 31) | 9  (3, 19) | 5  (1, 10) | 3  (1, 7) | 13  (5, 27) | 11  (4, 23) | 0.9  (0.2, 2.3) | 0.5  (0.1, 1.4) |
| 64,000-95,999 | 4.4  (0.20) | 2.7  (0.7, 6.7) | 25.0  (20.6, 33.5) | 474  (215, 830) | 299  (133, 528) | 19  (7, 36) | 10  (3, 20) | 6  (2, 11) | 3  (1, 8) | 13  (6, 26) | 10  (3, 23) | 1.2  (0.3, 2.4) | 0.6  (0.1, 1.6) |
| >96,000 | 4.5  (0.23) | 4.5  (2.7, 6.7) | 25.5  (20.2, 31.1) | 482  (281, 766) | 264  (111, 446) | 21  (10, 32) | 9  (3, 17) | 8  (5, 12) | 3  (1, 6) | 10  (7, 18) | 7  (2, 13) | 1.2  (0.5, 2.2) | 0.5  (0.1, 1.3) |
| **Highest education level** |  |  |  |  |  |  |  |  |  |  |  |  |  |
| No or compulsory school-level qualifications (basic) | 4.4  (0.05) | 2.0  (0.0, 4.5) | 27.3  (21.5, 33.6) | 466  (228, 828) | 292  (129, 509) | 17  (7, 35) | 9  (2, 20) | 6  (2, 11) | 3  (1, 7) | 16  (6, 30) | 11  (4, 23) | 1.2  (0.3, 2.5) | 0.5  (0.1, 1.5) |
| University entry qualifications and vocational equivalents (further) | 4.4  (0.11) | 2.1  (0.0, 5.0) | 26.8  (18.7, 31.9) | 452  (233, 801) | 289  (126, 504) | 18  (6, 32) | 9  (2, 19) | 6  (2, 12) | 3  (1, 7) | 13  (5, 28) | 10  (3, 23) | 1.4  (0.3, 2.5) | 0.5  (0.1, 1.4) |
| Degree level qualifications (degree). | 4.5  (0.05) | 2.5  (0.0, 5.4) | 26.7  (21.0, 33.8) | 424  (209, 765) | 296  (129, 504) | 16  (6, 32) | 9  (3, 20) | 5  (2, 10) | 3  (1, 7) | 13  (5, 28) | 11  (4, 23) | 1.0  (0.3, 2.3) | 0.5  (0.1, 1.5) |
| **IMD quintile** |  |  |  |  |  |  |  |  |  |  |  |  |  |
| 1 (most deprived) | 4.4  (0.09) | 2.0  (0.0, 5.0) | 25.8  (20.2, 31.9) | 468  (233, 828) | 297  (139, 497) | 20  (8, 35) | 9  (3, 19) | 6  (2, 12) | 3  (1, 7) | 14  (4, 29) | 11  (4, 23) | 1.2  (0.3, 2.6) | 0.5  (0.1, 1.5) |
| 2 | 4.3  (0.08) | 2.5  (0.9, 5.1) | 25.9  (20.0, 32.5) | 473  (263, 854) | 308  (138, 513) | 19  (8, 35) | 10  (3, 20) | 6  (2, 11) | 3  (1, 7) | 14  (6, 32) | 11  (4, 24) | 1.3  (0.4, 2.5) | 0.5  (0.1, 1.5) |
| 3 | 4.5  (0.07) | 2.0  (0.0, 5.6) | 27.0  (21.1, 33.7) | 394  (155, 751) | 297  (127, 518) | 14  (4, 31) | 10  (2, 21) | 5  (1, 10) | 3  (1, 8) | 13  (5, 29) | 11  (4, 23) | 0.9  (0.2, 2.4) | 0.5  (0.1, 1.5) |
| 4 | 4.6  (0.08) | 2.5  (0.0, 5.2) | 28.2  (21.5, 34.1) | 452  (228, 734) | 278  (120, 488) | 16  (7, 32) | 9  (2, 18) | 5  (2, 10) | 3  (1, 7) | 14  (6, 28) | 10  (4, 22) | 1.1  (0.3, 2.3) | 0.5  (0.1, 1.4) |
| 5 (least deprived) | 4.6  (0.08) | 2.0  (0.0, 4.0) | 28.1  (22.6, 34.0) | 413  (218, 766) | 293  (126, 512) | 15  (6, 32) | 9  (3, 20) | 5  (2, 10) | 3  (1, 7) | 13  (5, 27) | 11  (4, 23) | 1.0  (0.3, 2.3) | 0.5  (0.1, 1.4) |
| **Urban/rural location** |  |  |  |  |  |  |  |  |  |  |  |  |  |
| Urban | 4.4  (0.04) | 2.2  (0.0, 5.3) | 26.1  (20.5, 33.6) | 444  (218, 789) | 295  (129, 507) | 17  (6, 33) | 9  (3, 20) | 5  (2, 11) | 3  (1, 7) | 14  (6, 29) | 11  (4, 23) | 1.1  (0.3, 2.4) | 0.5  (0.1, 1.5) |
| Rural | 4.6  (0.08) | 2.0  (0.0, 3.8) | 29.5  (23.9, 33.8) | 424  (207, 751) | 290  (126, 493) | 16  (5, 32) | 9  (2, 19) | 5  (2, 10) | 3  (1, 7) | 14  (5, 28) | 11  (4, 23) | 0.8  (0.2, 2.4) | 0.5  (0.1, 1.4) |
| **Economic activity** |  |  |  |  |  |  |  |  |  |  |  |  |  |
| Active | 4.4  (0.04) | 2.3  (0.0, 5.2) | 26  (20.3, 32.9) | 446  (211, 783) | 305  (140, 515) | 17  (6, 32) | 10  (3, 20) | 5  (2, 11) | 3  (1, 7) | 14  (5, 29) | 11  (4, 24) | 1.1  (0.3, 2.4) | 0.5  (0.1, 1.5) |
| Inactive | 4.7  (0.08) | 2.0  (0.0, 4.5) | 29.3  (24.0, 35.4) | 431  (233, 775) | 264  (97, 473) | 16  (7, 33) | 8  (2, 18) | 6  (2, 12) | 3  (1, 7) | 13  (6, 28) | 9  (3, 20) | 1.1  (0.3, 2.3) | 0.4  (0.1, 1.3) |
| **Simplified NS-SEC analytic class** |  |  |  |  |  |  |  |  |  |  |  |  |  |
| 1 (Higher managerial, administrative and professional) | 4.5  (0.05) | 2.3  (0.0, 5.0) | 26.7  (21.0, 33.7) | 435  (215, 777) | 297  (131, 505) | 18  (6, 34) | 9  (3, 20) | 5  (2, 11) | 3  (1, 7) | 13  (5, 28) | 11  (4, 23) | 1.1  (0.3, 2.4) | 0.5  (0.1, 1.5) |
| 2 (intermediate) | 4.5  (0.07) | 2.0  (0.0, 5.2) | 28.3  (21.0, 34.0) | 448  (227, 779) | 288  (124, 492) | 16  (7, 30) | 9  (2, 19) | 5  (2, 11) | 3  (1, 7) | 16  (6, 29) | 10  (4, 22) | 1.3  (0.3, 2.4) | 0.5  (0.1, 1.4) |
| 3 (routine or manual) | 4.5  (0.08) | 2.3  (0.0, 5.0) | 26.6  (20.8, 32.5) | 430  (198, 791) | 295  (132, 521) | 15  (5, 33) | 9  (2, 19) | 5  (1, 11) | 3  (1, 7) | 15  (5, 30) | 11  (4, 24) | 1.0  (0.2, 2.4) | 0.5  (0.1, 1.5) |
| Not classifiable | 4.3  (0.19) | 1.5  (0.0, 5.0) | 25.5  (20.2, 33.8) | 489  (275, 813) | 299  (112, 508) | 19  (8, 35) | 10  (2, 21) | 5  (2, 11) | 3  (1, 7) | 14  (5, 28) | 10  (4, 22) | 1.4  (0.5, 2.7) | 0.5  (0.1, 1.5) |

OOH – out-of-home; BMI – Body Mass Index; IMD – Index of Multiple Deprivation; NS-SEC – National Statistics Socio-Economic Classification based on occupation. a – weighted by weekday/weekend to account for potential differences in OOH use with time of week.

**Supplementary Table S8**. Population weighted median (Q1, Q3) energy, total fat, saturated fat and sugar intakes as a percentage of eating occasion energy (%E) from OOH eating occasions (n=2,837) and non-OOH eating occasions (n=21,345) by demographic characteristics for participants included in Models 1.0-1.8 and 2.0-2.8 (n=1232).

|  | **Eating occasion energy**  **(kcal)** | | **Total fat**  **(%E)** | | **Total saturated fat (%E)** | | **Total sugars**  **(%E)** | |
| --- | --- | --- | --- | --- | --- | --- | --- | --- |
|  | **OOH** | **non-OOH** | **OOH** | **non-OOH** | **OOH** | **non-OOH** | **OOH** | **non-OOH** |
| **All** | 440 (215, 783) | 294 (129, 505) | 34 (23, 43) | 30 (17, 41) | 11 (5, 17) | 10 (4, 17) | 13 (5, 28) | 17 (5, 36) |
| **Sex** |  |  |  |  |  |  |  |  |
| Female | 407 (209, 739) | 267 (118, 458) | 35 (25, 43) | 30 (17, 41) | 12 (5, 17) | 10 (4, 17) | 14 (5, 29) | 18 (6, 37) |
| Male | 477 (230, 843) | 331 (149, 560) | 33 (22, 43) | 30 (16, 40) | 10 (4, 16) | 10 (4, 16) | 12 (5, 27) | 16 (5, 36) |
| **Ethnicity** |  |  |  |  |  |  |  |  |
| White | 442 (215, 786) | 294 (128, 505) | 34 (23, 43) | 30 (17, 41) | 11 (5, 17) | 10 (4, 17) | 13 (5, 28) | 17 (5, 36) |
| Asian, Black, Mixed and Other | 410 (217, 726) | 306 (149, 507) | 34 (24, 42) | 31 (17, 43) | 10 (5, 15) | 9 (4, 16) | 13 (5, 29) | 15 (4, 33) |
| **Age (years)** |  |  |  |  |  |  |  |  |
| 18-29 | 488 (250, 856) | 489 (275, 813) | 36 (26, 42) | 31 (19, 42) | 11 (5, 17) | 10 (4, 16) | 14 (5, 29) | 16 (5, 35) |
| 30-44 | 472 (238, 832) | 308 (151, 518) | 36 (26, 44) | 31 (18, 42) | 11 (5, 17) | 10 (4, 17) | 10 (4, 23) | 16 (5, 35) |
| 45-59 | 438 (205, 779) | 284 (119, 501) | 33 (21, 41) | 29 (14, 40) | 11 (5, 16) | 10 (4, 16) | 13 (5, 30) | 18 (5, 37) |
| 60+ | 377 (189, 683) | 264 (103, 457) | 33 (21, 43) | 30 (16, 40) | 10 (4, 16) | 10 (4, 17) | 16 (5, 30) | 18 (6, 37) |
| **BMI (kg/m^2^)** |  |  |  |  |  |  |  |  |
| <25 | 427 (207, 771) | 294 (134, 503) | 34 (23, 43) | 31 (17, 41) | 11 (5, 17) | 10 (4, 17) | 14 (5, 29) | 18 (6, 36) |
| 25-29.9 | 432 (214, 759) | 288 (120, 498) | 33 (22, 42) | 30 (16, 40) | 11 (5, 17) | 10 (4, 17) | 12 (5, 29) | 17 (5, 37) |
| ≥30 | 483 (234, 824) | 306 (131, 518) | 36 (25, 44) | 30 (16, 41) | 11 (5, 17) | 10 (4, 17) | 12 (5, 26) | 16 (5, 36) |
| **Marital status** |  |  |  |  |  |  |  |  |
| Married, cohabiting or in civil partnership | 427 (214, 766) | 290 (128, 498) | 34 (23, 43) | 30 (17, 40) | 11 (5, 17) | 10 (4, 17) | 13 (5, 29) | 17 (5, 36) |
| Single, widowed or divorced | 464 (218, 828) | 305 (131, 521) | 34 (24, 42) | 31 (16, 42) | 10 (4, 16) | 10 (4, 17) | 14 (5, 27) | 17 (5, 36) |
| **Child status** |  |  |  |  |  |  |  |  |
| Has child/children living at home | 477 (229, 844) | 302 (133, 507) | 35 (25, 43) | 30 (17, 41) | 11 (5, 17) | 10 (4, 17) | 13 (5, 27) | 16 (5, 35) |
| No child/children living at home | 424 (212, 752) | 291 (126, 503) | 34 (23, 43) | 30 (16, 40) | 11 (5, 17) | 10 (4, 17) | 13 (5, 29) | 17 (5, 37) |
| **Equivalised household income (£)** |  |  |  |  |  |  |  |  |
| <19,000 | 446 (245, 761) | 293 (120, 515) | 34 (24, 43) | 30 (16, 41) | 11 (5, 17) | 10 (4, 17) | 15 (5, 29) | 17 (5, 36) |
| 19,000-31,999 | 471 (226, 822) | 297 (134, 513) | 35 (24, 43) | 30 (17, 41) | 11 (6, 17) | 10 (4, 17) | 13 (4, 28) | 18 (6, 37) |
| 32,000-63,999 | 403 (195, 765) | 292 (134, 488) | 33 (22, 42) | 30 (16, 40) | 10 (4, 16) | 10 (4, 16) | 12 (5, 29) | 17 (5, 36) |
| 64,000-95,999 | 474 (215, 830) | 299 (133, 528) | 36 (25, 43) | 33 (19, 42) | 11 (6, 17) | 11 (5, 19) | 12 (5, 22) | 16 (5, 33) |
| >96,000 | 482 (281, 766) | 264 (111, 446) | 37 (31, 45) | 31 (16, 43) | 15 (13, 17) | 12 (4, 19) | 9 (5, 24) | 15 (5, 27) |
| **Highest education level** |  |  |  |  |  |  |  |  |
| No or compulsory school-level qualifications (basic) | 466 (228, 828) | 292 (129, 509) | 34 (24, 43) | 30 (16, 40) | 11 (5, 17) | 10 (4, 17) | 13 (5, 29) | 17 (5, 37) |
| University entry qualifications and vocational equivalents (further) | 452 (233, 801) | 289 (126, 504) | 34 (23, 43) | 29 (15, 40) | 11 (5, 16) | 9 (4, 16) | 10 (4, 25) | 15 (5, 36) |
| Degree level qualifications (degree). | 424 (209, 765) | 296 (129, 504) | 34 (23, 43) | 31 (18, 41) | 11 (5, 17) | 10 (4, 17) | 14 (5, 29) | 18 (5, 36) |
| **IMD quintile** |  |  |  |  |  |  |  |  |
| 1 (most deprived) | 468 (233, 828) | 297 (139, 497) | 37 (27, 45) | 30 (16, 41) | 12 (5, 17) | 10 (4, 16) | 12 (5, 27) | 16 (5, 35) |
| 2 | 473 (263, 854) | 308 (138, 513) | 34 (24, 42) | 30 (17, 41) | 11 (5, 16) | 11 (4, 17) | 12 (5, 28) | 17 (5, 36) |
| 3 | 394 (155, 751) | 297 (127, 518) | 33 (19, 42) | 31 (16, 41) | 10 (4, 16) | 10 (4, 17) | 14 (5, 29) | 17 (5, 37) |
| 4 | 452 (228, 734) | 278 (120, 488) | 34 (23, 42) | 30 (15, 39) | 10 (5, 17) | 9 (4, 16) | 13 (5, 27) | 17 (5, 36) |
| 5 (least deprived) | 413 (218, 766) | 293 (126, 512) | 34 (23, 43) | 31 (18, 41) | 11 (5, 17) | 10 (4, 17) | 14 (4, 30) | 18 (6, 36) |
| **Urban/rural location** |  |  |  |  |  |  |  |  |
| Urban | 444 (218, 789) | 295 (129, 507) | 34 (23, 43) | 30 (16, 41) | 11 (5, 17) | 10 (4, 17) | 13 (5, 28) | 17 (5, 36) |
| Rural | 424 (207, 751) | 290 (126, 493) | 34 (24, 43) | 30 (17, 40) | 11 (5, 18) | 10 (4, 17) | 14 (5, 32) | 18 (6, 37) |
| **Economic activity** |  |  |  |  |  |  |  |  |
| Active | 446 (211, 783) | 305 (140, 515) | 34 (24, 43) | 30 (17, 41) | 11 (5, 17) | 10 (4, 16) | 13 (5, 28) | 17 (5, 36) |
| Inactive | 431 (233, 775) | 264 (97, 473) | 34 (23, 43) | 30 (15, 40) | 12 (5, 17) | 10 (4, 17) | 12 (5, 28) | 18 (6, 36) |
| **Simplified NS-SEC analytic class** |  |  |  |  |  |  |  |  |
| 1 (Higher managerial, administrative and professional) | 435 (215, 777) | 297 (131, 505) | 34 (25, 43) | 31 (18, 41) | 11 (5, 17) | 10 (4, 17) | 13 (5, 27) | 17 (5, 36) |
| 2 (intermediate) | 448 (227, 779) | 288 (124, 492) | 34 (23, 42) | 30 (16, 40) | 11 (5, 17) | 10 (4, 17) | 14 (5, 29) | 18 (5, 37) |
| 3 (routine or manual) | 430 (198, 791) | 295 (132, 521) | 34 (21, 44) | 29 (14, 40) | 10 (4, 16) | 10 (4, 16) | 14 (5, 30) | 17 (5, 37) |
| Not classifiable | 489 (275, 813) | 299 (112, 508) | 34 (23, 43) | 31 (17, 43) | 8 (4, 15) | 10 (4, 16) | 11 (4, 20) | 15 (5, 34) |

OOH – out-of-home; BMI – Body Mass Index; IMD – Index of Multiple Deprivation; NS-SEC – National Statistics Socio-Economic Classification based on occupation.

**Supplementary Table S9**. Population weighted median (Q1, Q3) energy, total fat, saturated fat and sugar intakes as a percentage of daily energy (%E) from OOH days (n=1,897) and non-OOH days (n=3,545) by demographic characteristics for participants included in Models 1.0-1.8 and 2.0-2.8 (N=1232).

|  | **Daily energy**  **(kcal)** | | **Total fat**  **(%E)** | | **Total saturated fat**  **(%E)** | | **Total sugars**  **(%E)** | |
| --- | --- | --- | --- | --- | --- | --- | --- | --- |
|  | **OOH** | **non-OOH** | **OOH** | **non-OOH** | **OOH** | **non-OOH** | **OOH** | **non-OOH** |
| **All** | 1774 (1390, 2262) | 1575 (1188, 1989) | 34 (29, 40) | 33 (28, 39) | 12 (9, 15) | 12 (9, 15) | 16 (11, 21) | 16 (11, 22) |
| **Sex** |  |  |  |  |  |  |  |  |
| Female | 1659 (1276, 2067) | 1462 (1120, 1834) | 34 (30, 40) | 34 (28, 39) | 12 (10, 15) | 12 (9, 15) | 16 (11, 22) | 17 (11, 22) |
| Male | 1923 (1533, 2459) | 1680 (1265, 2137) | 34 (29, 39) | 33 (28, 39) | 12 (9, 15) | 12 (9, 15) | 15 (10, 21) | 15 (10, 21) |
| **Ethnicity** |  |  |  |  |  |  |  |  |
| White | 1772 (1393, 2260) | 1582 (1189, 1991) | 34 (29, 40) | 33 (28, 39) | 12 (9, 15) | 12 (9, 15) | 16 (11, 21) | 16 (11, 22) |
| Asian, Black, Mixed and Other | 1793 (1262, 2319) | 1422 (1051, 1862) | 35 (30, 39) | 33 (28, 40) | 11 (9, 14) | 11 (8, 14) | 16 (10, 21) | 16 (10, 21) |
| **Age (years)** |  |  |  |  |  |  |  |  |
| 18-29 | 1777 (1438, 2305) | 1584 (1194, 1997) | 35 (30, 40) | 33 (28, 40) | 12 (9, 15) | 12 (9, 15) | 16 (11, 22) | 16 (11, 22) |
| 30-44 | 1784 (1368, 2285) | 1611 (1213, 2046) | 35 (30, 40) | 34 (29, 39) | 12 (10, 15) | 12 (9, 15) | 15 (10, 20) | 16 (11, 22) |
| 45-59 | 1835 (1415, 2317) | 1579 (1163, 2001) | 33 (28, 39) | 33 (28, 39) | 12 (9, 15) | 12 (9, 15) | 16 (11, 22) | 15 (10, 21) |
| 60+ | 1721 (1318, 2140) | 1481 (1132, 1835) | 33 (28, 39) | 33 (28, 38) | 12 (9, 15) | 12 (10, 15) | 16 (11, 22) | 16 (11, 22) |
| **BMI (kg/m^2^)** |  |  |  |  |  |  |  |  |
| <25 | 1765 (1409, 2218) | 1596 (1215, 1999) | 34 (29, 40) | 33 (28, 39) | 12 (9, 15) | 12 (9, 15) | 16 (11, 22) | 17 (12, 23) |
| 25-29.9 | 1831 (1390, 2343) | 1550 (1143, 1947) | 33 (29, 39) | 33 (28, 39) | 12 (9, 15) | 12 (9, 15) | 16 (10, 22) | 15 (10, 22) |
| ≥30 | 1736 (1342, 2229) | 1568 (1182, 1977) | 35 (30, 40) | 34 (29, 39) | 12 (10, 15) | 12 (9, 16) | 15 (10, 21) | 15 (10, 20) |
| **Marital status** |  |  |  |  |  |  |  |  |
| Married, cohabiting or in civil partnership | 1763 (1376, 2245) | 1571 (1199, 1983) | 34 (29, 39) | 33 (28, 39) | 12 (9, 15) | 12 (9, 15) | 16 (10, 22) | 16 (11, 21) |
| Single, widowed or divorced | 1819 (1404, 2309) | 1587 (1162, 1998) | 35 (30, 40) | 34 (28, 40) | 12 (9, 15) | 12 (9, 15) | 16 (11, 21) | 17 (11, 22) |
| **Child status** |  |  |  |  |  |  |  |  |
| Has child/children living at home | 1847 (1439, 2401) | 1583 (1225, 2022) | 34 (30, 39) | 34 (29, 39) | 12 (10, 15) | 12 (9, 15) | 15 (11, 21) | 15 (11, 21) |
| No child/children living at home | 1747 (1360, 2207) | 1570 (1163, 1962) | 34 (29, 40) | 33 (28, 39) | 12 (9, 15) | 12 (9, 15) | 16 (11, 22) | 16 (11, 22) |
| **Equivalised household income (£)** |  |  |  |  |  |  |  |  |
| <19,000 | 1772 (1396, 2319) | 1579 (1182, 2003) | 35 (29, 40) | 33 (28, 39) | 12 (9, 16) | 12 (9, 15) | 16 (11, 22) | 16 (11, 22) |
| 19,000-31,999 | 1772 (1368, 2254) | 1610 (1200, 1991) | 35 (30, 40) | 33 (28, 39) | 12 (9, 15) | 12 (9, 15) | 15 (10, 21) | 16 (11, 22) |
| 32,000-63,999 | 1775 (1395, 2178) | 1527 (1156, 1943) | 33 (28, 39) | 33 (27, 39) | 11 (9, 15) | 12 (8, 15) | 16 (12, 21) | 16 (11, 22) |
| 64,000-95,999 | 1908 (1500, 2630) | 1623 (1272, 2034) | 35 (31, 40) | 34 (29, 39) | 12 (10, 14) | 12 (9, 15) | 13 (10, 18) | 14 (9, 22) |
| >96,000 | 1435 (1188, 1737) | 1502 (1378, 1721) | 34 (32, 37) | 32 (21, 35) | 14 (11, 15) | 11 (8, 15) | 14 (8, 17) | 11 (6, 16) |
| **Highest education level** |  |  |  |  |  |  |  |  |
| No or compulsory school-level qualifications (basic) | 1816 (1385, 2276) | 1547 (1158, 1969) | 35 (29, 40) | 33 (28, 38) | 12 (9, 15) | 12 (9, 15) | 16 (10, 22) | 16 (10, 22) |
| University entry qualifications and vocational equivalents (further) | 1673 (1283, 2200) | 1553 (1169, 1965) | 35 (29, 39) | 32 (26, 38) | 12 (9, 15) | 12 (8, 15) | 15 (10, 21) | 15 (11, 21) |
| Degree level qualifications (degree). | 1788 (1403, 2262) | 1596 (1200, 2001) | 34 (29, 40) | 34 (28, 39) | 12 (9, 15) | 12 (9, 15) | 16 (11, 21) | 16 (11, 22) |
| **IMD quintile** |  |  |  |  |  |  |  |  |
| 1 (most deprived) | 1731 (1307, 2361) | 1533 (1151, 1977) | 35 (29, 40) | 33 (28, 39) | 12 (10, 16) | 12 (9, 15) | 15 (10, 20) | 15 (10, 21) |
| 2 | 1849 (1385, 2309) | 1566 (1173, 1991) | 35 (29, 39) | 33 (28, 39) | 12 (9, 15) | 12 (9, 15) | 15 (11, 22) | 16 (11, 22) |
| 3 | 1723 (1308, 2159) | 1630 (1223, 2043) | 34 (29, 40) | 34 (29, 40) | 12 (9, 15) | 13 (10, 16) | 17 (11, 22) | 15 (11, 22) |
| 4 | 1784 (1455, 2242) | 1518 (1145, 1903) | 33 (29, 38) | 32 (27, 38) | 11 (9, 15) | 11 (8, 15) | 16 (11, 22) | 16 (10, 21) |
| 5 (least deprived) | 1837 (1423, 2305) | 1628 (1219, 2001) | 34 (29, 40) | 34 (29, 39) | 12 (10, 15) | 12 (9, 15) | 15 (11, 21) | 17 (11, 23) |
| **Urban/rural location** |  |  |  |  |  |  |  |  |
| Urban | 1772 (1395, 2257) | 1569 (1182, 1983) | 34 (29, 40) | 33 (28, 39) | 12 (9, 15) | 12 (9, 15) | 16 (11, 21) | 16 (11, 22) |
| Rural | 1779 (1362, 2297) | 1606 (1199, 1997) | 34 (28, 40) | 33 (27, 39) | 12 (9, 16) | 12 (9, 15) | 17 (11, 23) | 16 (11, 22) |
| **Economic activity** |  |  |  |  |  |  |  |  |
| Active | 1793 (1402, 2305) | 1597 (1197, 2003) | 34 (29, 40) | 33 (28, 39) | 12 (9, 15) | 12 (9, 15) | 16 (11, 22) | 16 (11, 22) |
| Inactive | 1728 (1308, 2142) | 1508 (1142, 1901) | 34 (29, 39) | 34 (28, 39) | 12 (9, 16) | 12 (9, 15) | 15 (10, 21) | 16 (11, 21) |
| **Simplified NS-SEC analytic class** |  |  |  |  |  |  |  |  |
| 1 (Higher managerial, administrative and professional) | 1784 (1415, 2276) | 1601 (1200, 2001) | 35 (30, 39) | 34 (28, 39) | 12 (9, 15) | 12 (9, 15) | 16 (11, 21) | 16 (11, 22) |
| 2 (intermediate) | 1767 (1351, 2231) | 1571 (1134, 1951) | 33 (28, 39) | 34 (28, 39) | 12 (9, 16) | 12 (9, 15) | 16 (11, 21) | 15 (11, 22) |
| 3 (routine or manual) | 1801 (1374, 2305) | 1558 (1176, 2002) | 34 (28, 40) | 32 (26, 38) | 12 (9, 15) | 12 (9, 15) | 16 (11, 24) | 16 (11, 22) |
| Not classifiable | 1599 (1290, 2007) | 1505 (1181, 1877) | 34 (29, 40) | 35 (29, 40) | 12 (9, 15) | 12 (8, 14) | 13 (8, 21) | 16 (11, 22) |

OOH – out-of-home; BMI – Body Mass Index; IMD – Index of Multiple Deprivation; NS-SEC – National Statistics Socio-Economic Classification based on occupation.

**Supplementary Table S10**. Effect estimates from eating occasion-level models 1.0-1.8 and day-level models 2.0-2.8 (N=1232).

|  | **Models 1.0-1.8**  **Eating occasion-level energy** | | | | | **Models 2.0-2.8 Day-level energy** | | | |
| --- | --- | --- | --- | --- | --- | --- | --- | --- | --- |
|  | **β** | **95% CI** | **p-value** | **Adjusted p-value ^c^** | **Β** | | **95% CI** | **p-value** | **Adjusted p-value ^c^** |
| **Model .0 - unadjusted for potential confounders ^a^** |  |  |  |  |  | |  |  |  |
| Non-OOH | *Ref=368* |  |  |  | *Ref=1641* | |  |  |  |
| OOH | 199 | 174, 224 | <0.001 * | 6.64x10^-53^ | 111 | | 38, 183 | 0.003 * | 0.039 |
| **Model .1 - adjusted for potential confounders ^b^** |  |  |  |  |  | |  |  |  |
| Non-OOH | *Ref=377* |  |  |  | *Ref=1494* | |  |  |  |
| OOH | 196 | 171, 221 | <0.001 * | 5.63x10^-52^ | 103 | | 29, 177 | 0.006 | 0.063 |
| **Model .2 - Model .1 adjusted for age×OOH eating occasion or day** |  |  |  |  |  | |  |  |  |
| Age 18-29 | *Ref=226* |  |  |  | *Ref=161* | |  |  |  |
| Age 30-44 | -9 | -85, 67 | 0.821 | 1.000 | -27 | | -142, 88 | 0.646 | 1.000 |
| Age 45-59 | -41 | -112, 31 | 0.267 | 0.660 | -98 | | -206, 11 | 0.079 | 0.332 |
| Age 60+ | -62 | -128, 4 | 0.067 | 0.402 | -91 | | -197, 15 | 0.093 | 0.326 |
| **Model .3 - Model .1 adjusted for sex×OOH eating occasion or day** |  |  |  |  |  | |  |  |  |
| Female | *Ref=199* |  |  |  | *Ref=72* | |  |  |  |
| Male | -6 | -56, 44 | 0.816 | 1.000 | 68 | | -8, 144 | 0.079 | 0.302 |
| **Model .4 - Model .1 adjusted for BMI×OOH eating occasion or day** |  |  |  |  |  | |  |  |  |
| BMI <25 | *Ref=191* |  |  |  | *Ref=110* | |  |  |  |
| BMI 25-29.9 | -6 | -62, 50 | 0.837 | 1.000 | 13 | | -75, 102 | 0.765 | 1.000 |
| BMI ≥30 | 31 | -36, 98 | 0.359 | 0.754 | -44 | | -140, 51 | 0.362 | 0.724 |
| **Model .5 - Model .1 adjusted for education×OOH eating occasion or day** |  |  |  |  |  | |  |  |  |
| No or compulsory school-level qualifications (basic) | *Ref=222* |  |  |  | *Ref=156* | |  |  |  |
| University entry qualifications and vocational equivalents (further) | -23 | -110, 64 | 0.601 | 1.000 | -115 | | -252, 22 | 0.100 | 0.323 |
| Degree level qualifications (degree) | -41 | -91, 9 | 0.106 | 0.318 | -72 | | -148, 4 | 0.064 | 0.448 |
| **Model .6 - Model .1 adjusted for IMD×OOH eating occasion or day** |  |  |  |  |  | |  |  |  |
| IMD quintile 1 (most deprived) | *Ref=234* |  |  |  | *Ref=137* | |  |  |  |
| IMD quintile 2 | -2 | -82, 78 | 0.968 | 0.968 | -13 | | -139, 113 | 0.841 | 1.000 |
| IMD quintile 3 | -76 | -158, 7 | 0.073 | 0.383 | -110 | | -231, 11 | 0.076 | 0.355 |
| IMD quintile 4 | -51 | -125, 24 | 0.181 | 0.507 | -9 | | -130, 113 | 0.890 | 1.000 |
| IMD quintile 5 (least deprived) | -48 | -131, 34 | 0.253 | 0.664 | -36 | | -165, 93 | 0.583 | 1.000 |
| **Model .7 - Model .1 adjusted for occupational social class×OOH eating occasion or day** |  |  |  |  |  | |  |  |  |
| NS-SEC 1 (Higher managerial, administrative and professional) | *Ref=190* |  |  |  | *Ref=101* | |  |  |  |
| NS-SEC 2 (Intermediate) | 17 | -45, 80 | 0.586 | 1.000 | 16 | | -85, 117 | 0.751 | 1.000 |
| NS-SEC 3 (routine/manual) | -2 | -65, 62 | 0.960 | 0.983 | 5 | | -87, 97 | 0.913 | 1.000 |
| Not classifiable | 55 | -46, 157 | 0.287 | 0.670 | -55 | | -187, 77 | 0.416 | 0.794 |
| **Model .8 - Model .1 adjusted for equivalised household income×OOH eating occasion or day** |  |  |  |  |  | |  |  |  |
| <£19,000 | *Ref=197* |  |  |  | *Ref=104* | |  |  |  |
| £19,000-£31,999 | -2 | -62, 57 | 0.943 | 1.000 | -7 | | -100, 87 | 0.886 | 1.000 |
| £32,000-£63,999 | -5 | -68, 57 | 0.867 | 1.000 | -3 | | -96, 91 | 0.955 | 1.000 |
| £64,000-£95,999 | 34 | -97, 165 | 0.613 | 0.990 | 134 | | -115, 383 | 0.291 | 0.643 |
| >£96,000 | 27 | -128, 183 | 0.731 | 1.000 | -225 | | -462, 11 | 0.062 | 0.521 |

OOH – out-of-home; BMI – Body Mass Index; IMD – Index of Multiple Deprivation; NS-SEC – National Statistics Socio-Economic Classification; Ref – reference category value

Reference category in models 1.0 and 2.0 is the kcal for a non-OOH eating occasion/day, unadjusted for potential confounders. Reference category in models 1.1 and 2.1 is the kcal for a non-OOH eating occasion/day for someone aged 18-29, female, BMI<25kg/m^2^, no or compulsory school-level qualifications, IMD quintile=1, NS-SEC category 1 (Higher managerial, administrative and professional), and <£19,000 equivalised household income. Reference category in models 1.2-1.8 and models 2.2-2.8 is the additional kcal for an OOH eating occasion/day compared with a non-OOH eating occasion/day for someone aged 18-29, female, BMI<25kg/m^2^, no or compulsory school-level qualifications, IMD quintile=1, NS-SEC category 1 (Higher managerial, administrative and professional), and <£19,000 equivalised household income.

β coefficients in models 1.2-1.8 and models 2.2-2.8 represent the difference in energy intakes between an OOH eating occasion/day and non-OOH eating occasion/day for a given sub-group over and above the reference sub-group intake. In all models, sample data was weighted to be representative of the 2021 England Census adult population sub-group sizes for age, sex, ethnicity and highest education level.

a – Model 1.0 is conducted in participants with complete covariate data. Model 2.0 is adjusted for frequency per day of OOH eating occasions and conducted in participants with complete covariate data.

b - Model 1.1 and 2.1 are adjusted for age, sex, BMI, ethnicity, having a child <18 years living at home, highest level of education, IMD, urban/rural location, occupational social class (NS-SEC), equivalised household income, time of year/season and week/weekend day. Model 2.1 is additionally adjusted for frequency per day of eating occasions containing OOH foods.

c - Benjamini-Hochberg adjusted p-values calculated as p-value*(number of tests/p-value rank)

* evidence of association after Benjamini-Hochberg adjustment for multiple testing.

**Supplementary Table S11**. Estimated energy misreporting status by demographic characteristic for participants included in Models 1.0-1.8 and 2.0-2.8.

|  | **Energy reporting status, n(%)** | | |
| --- | --- | --- | --- |
|  | **Under-reporting** | **Plausible-reporting** | **Over-reporting** |
| **All** | 791 (64%) | 386 (31%) | 55 (4%) |
| **Sex** |  |  |  |
| Female | 354 (57%) | 235 (38%) | 35 (6%) |
| Male | 437 (72%) | 151 (25%) | 20 (3%) |
| **Ethnicity** |  |  |  |
| White | 715 (65%) | 341 (31%) | 50 (5%) |
| Asian, Black, Mixed and Other | 76 (60%) | 45 (36%) | 5 (4%) |
| **Age (years)** |  |  |  |
| 18-29 | 210 (70%) | 81 (27%) | 11 (4%) |
| 30-44 | 208 (68%) | 89 (29%) | 9 (3%) |
| 45-59 | 186 (61%) | 105 (34%) | 16 (5%) |
| 60+ | 187 (59%) | 111 (35%) | 19 (6%) |
| **BMI (kg/m^2^)** |  |  |  |
| <25 | 285 (55%) | 201 (39%) | 35 (7%) |
| 25-29.9 | 277 (69%) | 110 (27%) | 14 (3%) |
| ≥30 | 229 (74%) | 75 (24%) | 6 (2%) |
| **Marital status** |  |  |  |
| Married, cohabiting or in civil partnership | 508 (64%) | 246 (31%) | 35 (4%) |
| Single, widowed or divorced | 283 (64%) | 140 (32%) | 20 (5%) |
| **Child status** |  |  |  |
| Has child/children living at home | 230 (60%) | 136 (36%) | 16 (4%) |
| No child/children living at home | 561 (66%) | 250 (29%) | 39 (5%) |
| **Equivalised household income (£)** |  |  |  |
| <19,000 | 296 (62%) | 163 (34%) | 22 (5%) |
| 19,000-31,999 | 241 (64%) | 118 (31%) | 17 (5%) |
| 32,000-63,999 | 224 (69%) | 90 (28%) | 13 (4%) |
| 64,000-95,999 | 23 (59%) | 13 (33%) | 3 (8%) |
| >96,000 | 7 (78%) | 2 (22%) | 0 (0%) |
| **Highest education level** |  |  |  |
| No or compulsory school-level qualifications (basic) | 393 (66%) | 183 (31%) | 24 (4%) |
| University entry qualifications and vocational equivalents (further) | 85 (65%) | 40 (31%) | 5 (4%) |
| Degree level qualifications (degree). | 313 (62%) | 163 (33%) | 25 (5%) |
| **IMD quintile** |  |  |  |
| 1 (most deprived) | 160 (71%) | 57 (25%) | 7 (3%) |
| 2 | 175 (64%) | 87 (32%) | 10 (4%) |
| 3 | 153 (61%) | 84 (34%) | 12 (5%) |
| 4 | 164 (63%) | 88 (34%) | 9 (3%) |
| 5 (least deprived) | 139 (62%) | 70 (31%) | 17 (8%) |
| **Urban/rural location** |  |  |  |
| Urban | 660 (65%) | 308 (30%) | 44 (4%) |
| Rural | 131 (60%) | 78 (35%) | 11 (5%) |
| **Economic activity** |  |  |  |
| Active | 592 (65%) | 281 (31%) | 33 (4%) |
| Inactive | 199 (61%) | 105 (32%) | 22 (7%) |
| **Simplified NS-SEC analytic class** |  |  |  |
| 1 (Higher managerial, administrative and professional) | 344 (64%) | 160 (30%) | 31 (6%) |
| 2 (intermediate) | 190 (62%) | 108 (35%) | 9 (3%) |
| 3 (routine or manual) | 199 (65%) | 92 (30%) | 14 (5%) |
| Not classifiable | 58 (68%) | 26 (31%) | 1 (1%) |

BMI – Body Mass Index; IMD – Index of Multiple Deprivation; NS-SEC – National Statistics Socio-Economic Classification based on occupation. Percentages may not add up to 100% due to rounding.

**Supplementary Table S12**. Sensitivity analysis – repeat of Models 1.1-1.8 and 2.1-2.8 (N=1232) with additional adjustment for energy misreporting status.

|  | **Models 1.1-1.8 Eating occasion-level energy** | | | | | **Models 2.1-2.8 Day-level energy** | | | |
| --- | --- | --- | --- | --- | --- | --- | --- | --- | --- |
| **Exposure** | **β** | **95% CI** | **p-value** | **Adjusted p-value ^b^** | **β** | | **95% CI** | **p-value** | **Adjusted p-value ^b^** |
| **Model .1 - adjusted for potential confounders ^a^** |  |  |  |  |  | |  |  |  |
| Non-OOH | *Ref=414* |  |  |  | *Ref=1808* | |  |  |  |
| OOH | 193 | 168, 218 | <0.001 * | 9.3x10^-50^ | 106 | | 34, 178 | 0.004 | 0.080 |
| **Model .2 - Model .1 adjusted for age×OOH eating occasion or day** |  |  |  |  |  | |  |  |  |
| Age 18-29 | *Ref=225* |  |  |  | *Ref=177* | |  |  |  |
| Age 30-44 | -12 | -88, 64 | 0.761 | 1.000 | -48 | | -155, 59 | 0.378 | 0.720 |
| Age 45-59 | -44 | -115, 27 | 0.226 | 0.565 | -106 | | -207, -5 | 0.039 | 0.312 |
| Age 60+ | -65 | -130, 1 | 0.053 | 0.303 | -114 | | -214, -14 | 0.025 | 0.333 |
| **Model .3 - Model .1 adjusted for sex×OOH eating occasion or day** |  |  |  |  |  | |  |  |  |
| Female | *Ref=195* |  |  |  | *Ref=69* | |  |  |  |
| Male | -5 | -56, 45 | 0.835 | 0.982 | 80 | | 10, 151 | 0.025 | 0.250 |
| **Model .4 - Model .1 adjusted for BMI×OOH eating occasion or day** |  |  |  |  |  | |  |  |  |
| BMI <25 | *Ref=189* |  |  |  | *Ref=115* | |  |  |  |
| BMI 25-29.9 | -9 | -65, 48 | 0.766 | 0.988 | 5 | | -77, 86 | 0.907 | 1.000 |
| BMI ≥30 | 31 | -36, 97 | 0.368 | 0.774 | -41 | | -130, 49 | 0.373 | 0.746 |
| **Model .5 - Model .1 adjusted for education×OOH eating occasion or day** |  |  |  |  |  | |  |  |  |
| No or compulsory school-level qualifications (basic) | *Ref=217* |  |  |  | *Ref=138* | |  |  |  |
| University entry qualifications and vocational equivalents (further) | -23 | -109, 64 | 0.606 | 0.970 | -85 | | -209, 39 | 0.178 | 0.593 |
| Degree level qualifications (degree) | -38 | -88, 12 | 0.136 | 0.544 | -38 | | -109, 33 | 0.292 | 0.649 |
| **Model .6 - Model .1 adjusted for IMD×OOH eating occasion or day** |  |  |  |  |  | |  |  |  |
| IMD quintile 1 (most deprived) | *Ref=230* |  |  |  | *Ref=157* | |  |  |  |
| IMD quintile 2 | 1 | -78, 80 | 0.971 | 0.971 | -26 | | -140, 89 | 0.661 | 1.000 |
| IMD quintile 3 | -76 | -159, 6 | 0.071 | 0.355 | -115 | | -227, -4 | 0.043 | 0.287 |
| IMD quintile 4 | -50 | -124, 24 | 0.185 | 0.569 | -42 | | -151, 67 | 0.449 | 0.816 |
| IMD quintile 5 (least deprived) | -52 | -135, 30 | 0.212 | 0.606 | -66 | | -182, 51 | 0.268 | 0.631 |
| **Model .7 - Model .1 adjusted for occupational social class×OOH eating occasion or day** |  |  |  |  |  | |  |  |  |
| NS-SEC 1 (Higher managerial, administrative and professional) | *Ref=187* |  |  |  | *Ref=100* | |  |  |  |
| NS-SEC 2 (Intermediate) | 17 | -45, 80 | 0.584 | 0.973 | 20 | | -72, 112 | 0.669 | 0.991 |
| NS-SEC 3 (routine and manual) | -2 | -65, 62 | 0.959 | 1.000 | 10 | | -75, 96 | 0.812 | 0.984 |
| NS-SEC 4 (student/unclassified) | 63 | -38, 163 | 0.224 | 0.597 | -26 | | -165, 112 | 0.710 | 0.979 |
| **Model .8 - Model .1 adjusted for equivalised household income×OOH eating occasion or day** |  |  |  |  |  | |  |  |  |
| <£19,000 | *Ref=191* |  |  |  | *Ref=97* | |  |  |  |
| £19,000-£31,999 | 1 | -58, 61 | 0.962 | 0.987 | 6 | | -81, 93 | 0.893 | 1.000 |
| £32,000-£63,999 | -4 | -66, 59 | 0.909 | 0.983 | 12 | | -73, 98 | 0.781 | 0.976 |
| £64,000-£95,999 | 40 | -91, 171 | 0.549 | 0.955 | 169 | | -61, 399 | 0.150 | 0.545 |
| >£96,000 | 32 | -126, 191 | 0.689 | 0.984 | -190 | | -422, 41 | 0.107 | 0.476 |

OOH – out-of-home; BMI – Body Mass Index; IMD – Index of Multiple Deprivation; NS-SEC – National Statistics Socio-Economic Classification; Ref – reference category value

Reference category in models 1.1 and 2.1 is the kcal for a non-OOH eating occasion/day for someone aged 18-29, female, BMI<25kg/m^2^, no or compulsory school-level qualifications, IMD quintile=1, NS-SEC category 1 (Higher managerial, administrative and professional), <£19,000 equivalised household income, and reporting plausible energy intakes. Reference category in models 1.2-1.8 and models 2.2-2.8 is the additional kcal for an OOH eating occasion/day compared with a non-OOH eating occasion/day for someone aged 18-29, female, BMI<25kg/m^2^, no or compulsory school-level qualifications, IMD quintile=1, NS-SEC category 1 (Higher managerial, administrative and professional), <£19,000 equivalised household income, and reporting plausible energy intakes.

β coefficients in models 1.2-1.8 and models 2.2-2.8 represent the difference in energy intakes between an OOH eating occasion/day and non-OOH eating occasion/day for a given sub-group over and above the reference sub-group intake. In all models, sample data was weighted to be representative of the 2021 England Census adult population sub-group sizes for age, sex, ethnicity and highest education level.

a - Model 1.1 and 2.1 are adjusted for age, sex, BMI, ethnicity, having a child <18 years living at home, highest level of education, IMD, urban/rural location, occupational social class (NS-SEC), equivalised household income, time of year/season, week/weekend day and misreporting status. Model 2.1 is additionally adjusted for frequency per day of eating occasions containing OOH foods.

b - Benjamini-Hochberg adjusted p-values calculated as p-value*(number of tests/p-value rank)

* evidence of association after Benjamini-Hochberg adjustment for multiple testing.

**Supplementary analyses specified in protocol**. To assess further the degree to which demographic covariates added between main models 1.0-1.1 and 2.0-2.1 contribute to explaining between- and within-person variation in energy intakes at the eating occasion- and day-level, models 1.0 and 2.0 were repeated with separate grouped adjustment for: 1) age, sex, BMI, ethnicity, and having a child <18 years living at home (model 1.0a and 2.0a); 2) education, IMD, urban/rural location, equivalised household income, and occupation (NS-SEC) (model 1.0b and 2.0b); and 3) time of year/season and week/weekend day (model 1.0c and 2.0c), and intraclass correlation coefficients calculated. Increases or decreases in the percentage of between-person variation explained between models 1.0 to 1.0a/1.0b/1.0c/1.1 or models 2.0 to 2.0a/2.0b/2.0c/2.1 indicate that the added covariates explain some of the differences in the outcome within or between individuals respectively.

| **Model** | **Adjustment variables** | **% between-person variation remaining in energy intake** |
| --- | --- | --- |
| **Eating occasion-level** |  |  |
| Model 1.0 | OOH/non-OOH (minimally adjusted) | 8.34 |
| Model 1.0a | OOH/non-OOH, age, sex, ethnicity, BMI, child/no child living at home | 6.29 |
| Model 1.0b | OOH/non-OOH, education, IMD, urban/rural location, equivalent household income, occupational class (NS-SEC) | 8.18 |
| Model 1.0c | OOH/non-OOH, time of year, time of week | 8.30 |
| Model 1.1 | OOH/non-OOH, age, sex, ethnicity, BMI, child/no child living at home, education, IMD, urban/rural location, equivalent household income, occupational social class (NS-SEC), time of year, time of week (fully adjusted) | 6.09 |
| **Day-level** |  |  |
| Model 2.0 | OOH/non-OOH, number of OOH eating occasions in day (minimally adjusted) | 42.54 |
| Model 2.0a | OOH/non-OOH, number of OOH eating occasions in day, age, sex, ethnicity, BMI, child/no child living at home | 39.62 |
| Model 2.0b | OOH/non-OOH, number of OOH eating occasions in day, education, IMD, urban/rural location, equivalent household income, occupational class (NS-SEC) | 42.21 |
| Model 2.0c | OOH/non-OOH, number of OOH eating occasions in day, time of year, time of week | 42.76 |
| Model 2.1 | OOH/non-OOH, number of OOH eating occasions in day, age, sex, ethnicity, BMI, child/no child living at home, education, IMD, urban/rural location, equivalent household income, occupational social class (NS-SEC), time of year, time of week (fully adjusted) | 39.52 |

OOH – out-of-home; BMI – Body Mass Index; IMD – Index of Multiple Deprivation; NS-SEC – National Statistics Socio-Economic Classification based on occupation.

10. Department for Environmental Food & Rural Affairs in collaboration with the Office for National Statistics. Guide to applying the Rural Urban Classification to data. 2016; July:11.

11. Office for National Statistics (ONS). A guide to labour market statistics. 2020. https://www.ons.gov.uk/employmentandlabourmarket/peopleinwork/employmentandemployeetypes/methodologies/aguidetolabourmarketstatistics. Accessed 23 Aug 2023.

12. Office for National Statistics (ONS). The National Statistics Socio-economic classification (NS-SEC). https://www.ons.gov.uk/methodology/classificationsandstandards/otherclassifications/thenationalstatisticssocioeconomicclassificationnssecrebasedonsoc2010. Accessed 23 Aug 2023.

13. Cornelsen L, Berger N, Cummins S, Smith RD. Socio-economic patterning of expenditures on ‘out-of-home’ food and non-alcoholic beverages by product and place of purchase in Britain. Soc Sci Med. 2019;235 June:112361.

14. Lake AA, Burgoine T, Greenhalgh F, Stamp E, Tyrrell R. The foodscape: Classification and field validation of secondary data sources. Heal Place. 2010;16:666–73.

15. Tyrrell RL, Greenhalgh F, Hodgson S, Wills WJ, Mathers JC, Adamson AJ, et al. Food environments of young people: Linking individual behaviour to environmental context. J Public Heal (United Kingdom). 2017;39:95–104.

16. Law C, Smith R, Cornelsen L. Place matters: Out-of-home demand for food and beverages in Great Britain. Food Policy. 2022;107 May 2021:102215.

17. Keeble M, Adams J, Burgoine T. Changes in Online Food Access during the COVID-19 Pandemic and Associations with Deprivation: Longitudinal Analysis. JMIR Public Heal Surveill. 2023;9.

18. World Health Organisation (WHO). The out-of-home food environment: report of a WHO Regional Office for Europe and Public Health England expert meeting, 10 June 2021. Copenhagen: WHO Regional Office for Europe; 2022.

19. Kalbus A, Cornelsen L, Ballatore A, Cummins S. Associations between the food environment and food and drink purchasing using large-scale commercial purchasing data: a cross-sectional study. BMC Public Health. 2023;23:1–15.

20. Food Standards Agency. Food portion sizes. 3rd edition. London: TSO; 2010.

21. Wrieden WL, Barton KL, Cochrane L, Adamson AJ. Calculation and collation of typical food portion sizes for adults aged 19-64 and older people aged 65 and over. Final Technical Report to the Food Standards Agency. 2006.

22. Office for National Statistics (ONS). Age, ethnic group, highest level of qualification and sex. Census 2021. 2021. https://www.ons.gov.uk/datasets/create/filter-outputs/1b461c9c-53a7-4288-aae0-793af87edb50. Accessed 10 Oct 2023.
